# Genomic Exploration of Essential Hypertension in African-Brazilian Quilombo Populations: A Comprehensive Approach with Pedigree Analysis and Family-Based Association Studies

**DOI:** 10.1101/2024.06.26.24309531

**Authors:** Vinícius Magalhães Borges, Andrea R.V.R. Horimoto, Ellen Marie Wijsman, Lilian Kimura, Kelly Nunes, Alejandro Q. Nato, Regina Célia Mingroni-Netto

## Abstract

**Background:** Essential Hypertension (EH) is a global health issue. Despite extensive research, much of EH heritability remains unexplained. We investigated the genetic basis of EH in African-derived individuals from partially isolated quilombo populations in Vale do Ribeira (SP-Brazil).

**Methods:** Samples from 431 individuals (167 affected, 261 unaffected, 3 unknown) were genotyped using a 650k SNP array. Estimated global ancestry proportions were 47% African, 36% European, and 16% Native American. We constructed six pedigrees using additional data from 673 individuals and created three non-overlapping SNP subpanels. We phased haplotypes and performed local ancestry analysis to account for admixture. Genome-wide linkage analysis (GWLA) and fine-mapping via family-based association studies (FBAS) were conducted, prioritizing EH-associated genes through systematic approach involving databases like PubMed, ClinVar, and GWAS Catalog.

**Results:** Linkage analysis identified 22 regions of interest (ROIs) with LOD scores ranging 1.45-3.03, encompassing 2,363 genes. Fine-mapping (FBAS) identified 60 EH-related candidate genes and 117 suggestive/significant variants. Among these, 14 genes, including *PHGDH*, *S100A10*, *MFN2*, and *RYR2*, were strongly related to hypertension harboring 29 suggestive/significant SNPs.

**Conclusions:** Through a complementary approach — combining admixture-adjusted GWLA based on Markov chain Monte Carlo methods, FBAS on known and imputed data, and gene prioritizing — new loci, variants, and candidate genes were identified. These findings provide targets for future research, replication in other populations, facilitate personalized treatments, and improve public health towards African-derived underrepresented populations. Limitations include restricted SNP coverage, self-reported pedigree data, and lack of available EH genomic studies on admixed populations for independent validation, despite the performed genetic correlation analyses using summary statistics.

**NOVELTY AND RELEVANCE:** *What Is New?:* This study applies a multi-level computational approach integrating admixture-adjusted genome-wide linkage analysis (GWLA), family-based association studies (FBAS), and fine-mapping strategies to investigate the genetic basis of essential hypertension (EH) in Brazilian Quilombo populations, a historically underrepresented group in genomic research.

*What Is Relevant?:* By focusing on admixed populations with high African ancestry, our findings address gaps in hypertension genetics by identifying 22 regions of interest (ROIs), 60 candidate genes, and 117 suggestive/significant variants, highlighting population-specific genetic factors that may contribute to EH risk. The study also emphasizes the need for ancestry-aware genomic analyses to improve the precision of genetic risk assessment in underrepresented populations.

*What Question Should Be Addressed Next?:* Future studies should focus on replicating these findings in independent admixed cohorts, conducting functional validation of prioritized genes, and integrating polygenic risk scores (PRS) adjusted for ancestry to enhance clinical applications for hypertension prevention and treatment in diverse populations.

## INTRODUCTION

Essential Hypertension (EH), OMIM: 145500, is a pervasive and sustained raise in arterial blood pressure (BP) and a major cause of premature death worldwide, responsible for approximately 9.4 million deaths annually.^1^ Classified as the primary preventable risk factor for cardiovascular diseases (CVDs),^2^ EH is defined as systolic BP (SBP) ≥ 140 mmHg and/or diastolic BP (DBP) ≥ 90 mmHg.^3–5^

EH, which affects 1.3 billion people worldwide annually,^6^ demands comprehensive investigation. Global prevalence is 33%,^6^ 23.9% in Brazil.^7^ EH is a multifactorial chronic condition, intricately weaving together environmental factors, social determinants, and greatly genetic/epigenetic influences, with 30-60% of estimated heritability.^8–10^ Its prevalence varies across different regions, affecting 35% of the population in the Americas, 28% in the Western Pacific, 37% in Europe, 32% in South-East Asia, 38% in the Eastern Mediterranean, and 36% in Africa.^6^ The risk for BP traits varies among ethnic groups^11^ and genetic ancestry significantly influences hypertension risk,^12^ particularly in African-derived populations.^1,13–16^ In the U.S., data shows that the prevalence of EH among non-Hispanic Black individuals is approximately 48.8%, notably higher than the prevalence among non-Hispanic White individuals at 37.6% and Hispanic individuals at 27.9%.^17^ Mortality rates due to EH and related diseases are also disproportionately higher, with African American individuals experiencing 4 to 5 times greater mortality than White adults.^18^

BP regulation involves complex interactions between the cardiovascular, renal, neural, and endocrine systems.^19^ The renin-angiotensin-aldosterone system (RAAS) plays a central role, with angiotensin II and aldosterone driving vasoconstriction and sodium retention.^10,20^ Baroreceptors and the sympathetic nervous system (SNS) provide rapid responses to BP changes,^21,22^ while endothelial cells release nitric oxide (NO) for vasodilation,^23^ The atrial natriuretic peptide (ANP) and the kallikrein-kinin system (KKS) counteract vasoconstriction and promote natriuresis.^24,25^ Sodium concentration also affects vascular tone through calcium exchange.^26^

The complex regulation is particularly relevant in populations as the African-derived.^27^ Genetic ancestry plays a crucial role in EH risk, with studies indicating that African-derived individuals generally develop higher BP starting in childhood. They excrete sodium more slowly and less completely than those of European descent, leading to volume-loading, which suppresses the RAAS and contributes to early-onset hypertension.^28,29^ Consequently, African-derived individuals often present a biochemical profile characterized by low or high plasma aldosterone, suppressed plasma renin activity or direct renin concentration, and reduced levels of angiotensin I and II.^30–32^ This results in a higher lifetime incidence of hypertension in African-derived populations compared with other populations.

Several genes have been found to be associated with an increased risk of EH specifically in African-derived populations, such as *ARMC5* (involved in RAAS^15^), *NOS3-GRK4* (involved in NO production and regulation^33^), *SCNN1B* and *SCNN1G* (involved in sodium channel^34^), *GRK4* (involved in sodium and water retention^13^), *SCG2* (confers regulation by PHOX2 transcription factors^35^), *AGT* (involved in angiotensin^36^) and *CYP11B2* (involved in alterations in aldosterone synthase production13).

Among social-environmental factors, EH risk is also influenced by adverse determinants of health, including overweight, smoking, physical inactivity,^9,37^ lower socioeconomic and educational status, concentrated poverty, and limited access to affordable, high-quality fresh food. Dietary patterns, alcohol consumption,^38^ particularly high-sodium^39–41^ and low-potassium intake, further elevate the risk.^42^ African-derived populations in lower social strata, such as those in the United States and South Africa, tend to experience higher rates of hypertension than expected based solely on anthropometric and socioeconomic factors.^43^

Yet, the genetic etiology of hypertension — encompassing genes, variants, susceptibility loci, and population disparities — remains elusive.^44,45^ Despite advancements from the common disease-common variant and common disease-rare variant hypotheses, methodologies such as Genome-Wide Linkage Analysis (GWLA) and Genome-Wide Association Studies (GWAS) face limitations.^46^ This gap leaves a segment of EH heritability unexplained by known genetic factors. Moreover, the existing underrepresentation (data as of December 2024) of African (0.19%), African American or Afro-Caribbean (0.45%), Hispanic or Latin American (0.34%), Other/Mixed (0.56%) populations^47^ in worldwide genomic investigations imposes constraints on the generalizability of results to admixed populations,^48,49^ such as the Brazilian one (68.1% European, 19.6% African, and 11.6% Native American).^50^

This study focuses on the tri-hybrid admixed populations known as “quilombo remnants” in Vale do Ribeira region, São Paulo, Brazil. Quilombo remnants are communities established by runaway or abandoned African enslaved individuals, often exhibiting intricate mixtures with European and Native American ancestry. These populations represent a unique model for the study of diseases. They are marked by a high prevalence of EH,^51^ well-defined clinical characterization, semi-isolation, background relatedness, high gene flow between populations, and founder effects.^52^ They also exhibit relatively homogeneous environmental influences, including lifestyle, dietary habits, and natural habitat, thereby minimizing confounding factors found in larger urban populations. Studying EH in quilombo remnants helps to reduce biases associated with population heterogeneity. This approach improves the signal-to-noise ratio, thereby enhancing statistical power, while providing better representation of admixed populations in genomic studies. It also allows us to uncover chromosomal regions, genes, and variants that may contribute to EH.

In this study, we employed a multi-level computational approach that combined both pedigree-based and population-based methodologies on family analysis to account for the unique admixture of African, European, and Native American ancestries in the quilombo population, along with a two-step fine-mapping strategy based on family-based association studies (FBAS) and investigation of EH-related genes. The family analysis using GWLA has been successfully applied in previous studies^53^ to address challenges such as population stratification and the complex genetic architecture of traits, enhancing the power to identify loci associated with rare variants that may go undetected in population-based studies. The pedigree- and population-based imputation methods we employed have also been rigorously developed and tested.^54,55^ FBAS enables detailed investigation of EH-related genes while controlling for population structure and relatedness, reducing the problem of multiple testing — an essential factor in admixed populations like the quilombo.^56,57^ Overall, by applying this strategy, we were able to fine-map candidate regions and variants associated with hypertension in a way that leverages the strengths of both family- and population-based genetic studies, providing insights into EH heritability in underrepresented populations.

## METHODS

Anonymized summary statistics data have been made publicly available at the GWAS Catalog (study GCST90454187) and can be accessed at https://www.ebi.ac.uk/gwas/studies/GCST90454187.

### SAMPLES AND SNP GENOTYPING

We conducted 51 trips to Vale do Ribeira (São Paulo, Brazil) from 2000-2020 to obtain samples (peripheral blood for DNA extraction), clinical data (average blood pressure, height, weight, waist circumference and hip circumference), and collect information (sex, age, family relationships, medical history, demographic information, and daily physical activity levels) from 431 consenting individuals aged 17 or older (Figure 1, box A). This study was approved by institutional review committees (USP/Institute of Biomedical Sciences 111/2001 and USP/Institute of Biosciences 012/2004 and 034/2005) and blood samples were drawn after subjects provided informed consent. To reduce bias, we excluded individuals who reported diabetes, severe kidney disease, and pregnancy. Additionally, no samples needed to be removed due to very low blood pressure (hypotensive readings), as no such cases were observed in the dataset.

**Figure 1.**
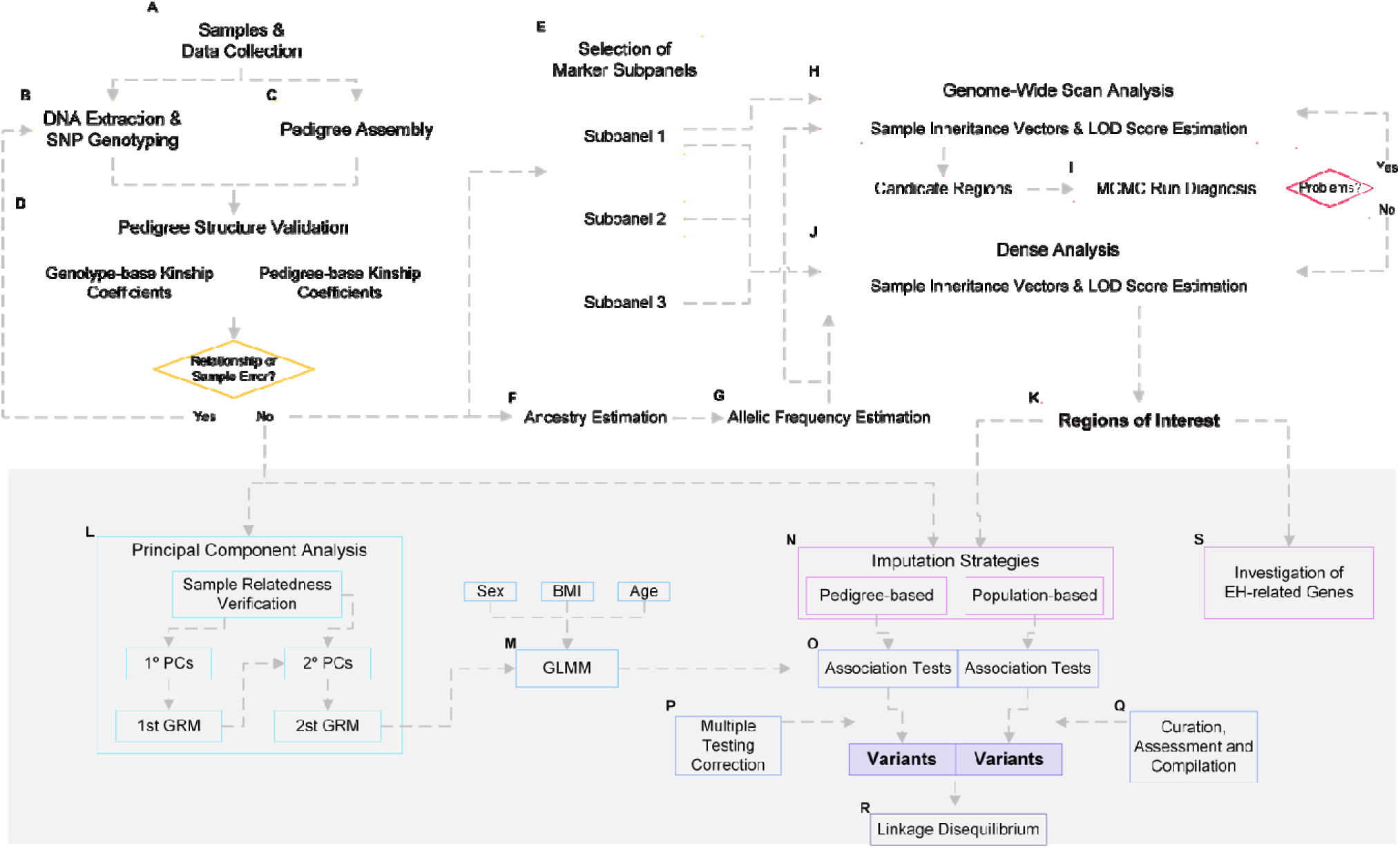
Schematic diagram of the main (boxes A-K) and fine-mapping (boxes L-S) strategy workflow. The color gradient progressively darkens to illustrate the advancement of the process. A) Sample collection and data preparation; B) DNA extraction, quantification, SNP genotyping, and quality control; C) Pedigree assembly; D) Pedigree structure validation; E) Marker selection for subpanels; F) Ancestry estimation; G) Allelic frequency estimation; H) Genome-wide scan analysis; I) Markov chain Monte Carlo (MCMC) convergence diagnostics; J) Dense mapping analysis; K) Identification of regions of interest (ROIs); L) Principal component analysis (PCA); M) Generalized linear mixed model fitting; N) Pedigree and population-based imputation; O) Family-based association tests; P) Multiple testing correction; Q) Variant curation and assessment; S) Investigation of EH-related genes.

Genomic DNA was extracted and quantified from each of the 431 blood samples and prepared for SNP genotyping through Axiom Genome-Wide Human Origins 1 Array SNPs (Figure 1, box B) according to Affymetrix requirements (details in Data S1). Raw data was processed, annotated, and subjected to quality control according to Affymetrix Human v.5a threshold^58^ using the commercial software Axiom Analysis Suite v.3.1.

From the combined collected data, we used the commercial software GenoPro v.3.0.1.4 – tool for creating and managing family trees and genealogical data – to construct six extended pedigrees from 8 different populations (Abobral [ABDR], André Lopes [AN], Galvão [GA], Ivaporunduva [IV], Nhunguara [NH], Pedro Cubas [PC], São Pedro [SP], and Sapatu [TU]), with detailed locations as previously described^59^ (Figure 2). The six pedigrees were named: **ABDR** (Abobral population); **ANNH** (André Lopes and Nhunguara populations); **GASP** (Galvão and São Pedro populations); **IV** (Ivaporunduva population); **PC** (Pedro Cubas population); and **TU** (Sapatu population). These pedigrees encompass 1,104 individuals (Table 1): 431 genotyped (167 affected, 261 non-affected and 3 unknown phenotype) and 673 non-genotyped. The characteristics of the genotyped cohort are detailed in Table 2. Pedigree structures underwent validation through calculation of multi-step pairwise kinship coefficients (*Φ*) using several algorithms: KING-Robust v.2.2.8^60^, which efficiently identifies relatedness in large datasets while accounting for population structure; MORGAN (Monte Carlo Genetic Analysis) v.3.4^61,62^, a robust suite designed for performing genetic linkage analysis in pedigrees; and PBAP (Pedigree-Based Analysis Pipeline) v.1/v.2^63^, a pipeline for file processing and quality control of pedigree data with dense genetic markers.

**Figure 2.**
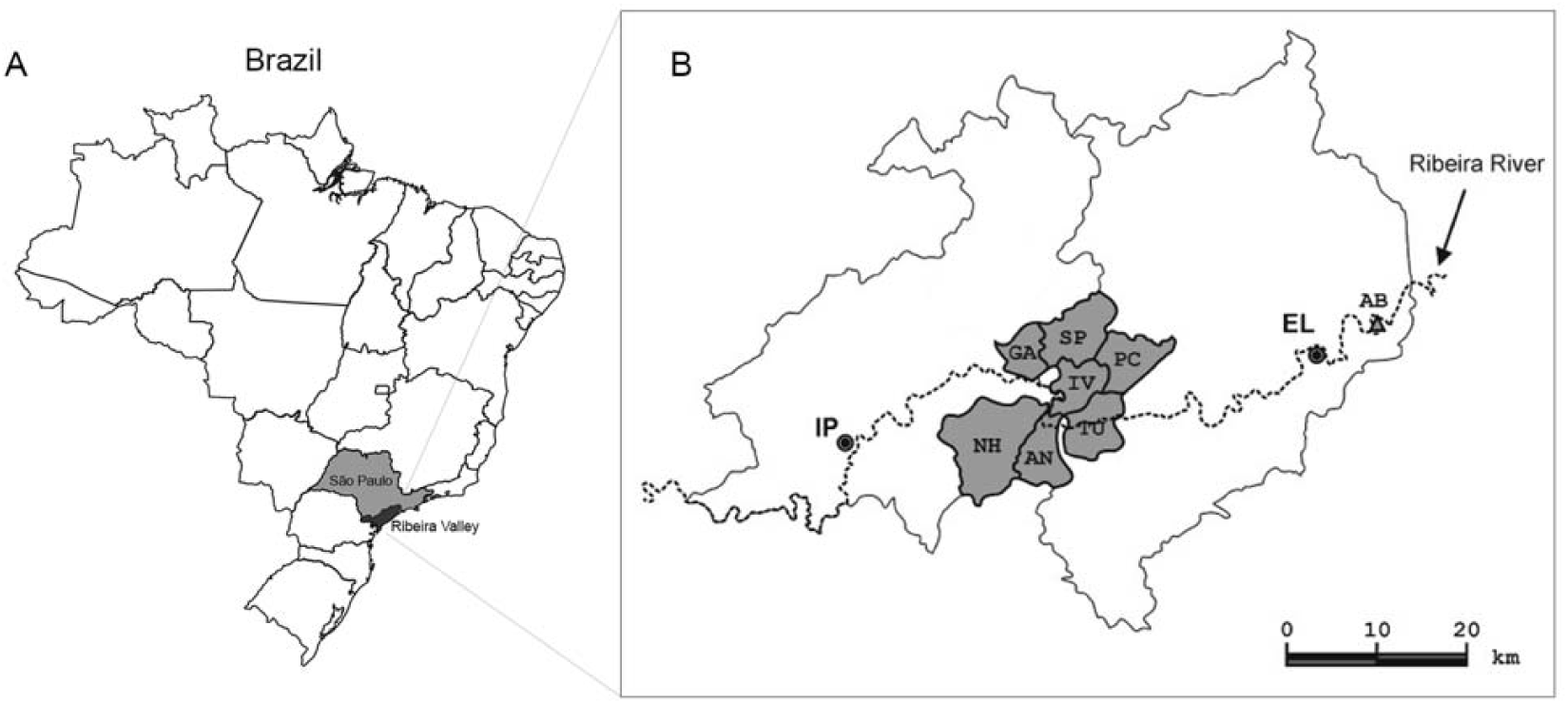
Geographical location of quilombo populations. A) Brazilian territory in South America, with the State of São Paulo highlighted in gray and the Ribeira Valley (*Vale do Ribeira*) in a darker shade of gray; B) Location of quilombo populations: AB (Abobral), AN (Andre Lopes), GA (Galvão), IV (Ivaporanduva), NH (Nhunguara), PC (Pedro Cubas), SP (São Pedro), and TU (Sapatu). The black dots denote the urban centers of Eldorado (EL) and Iporanga (IP). Adapted from Kimura L, *et al.* (2013) ^59^.

**Table 1.**
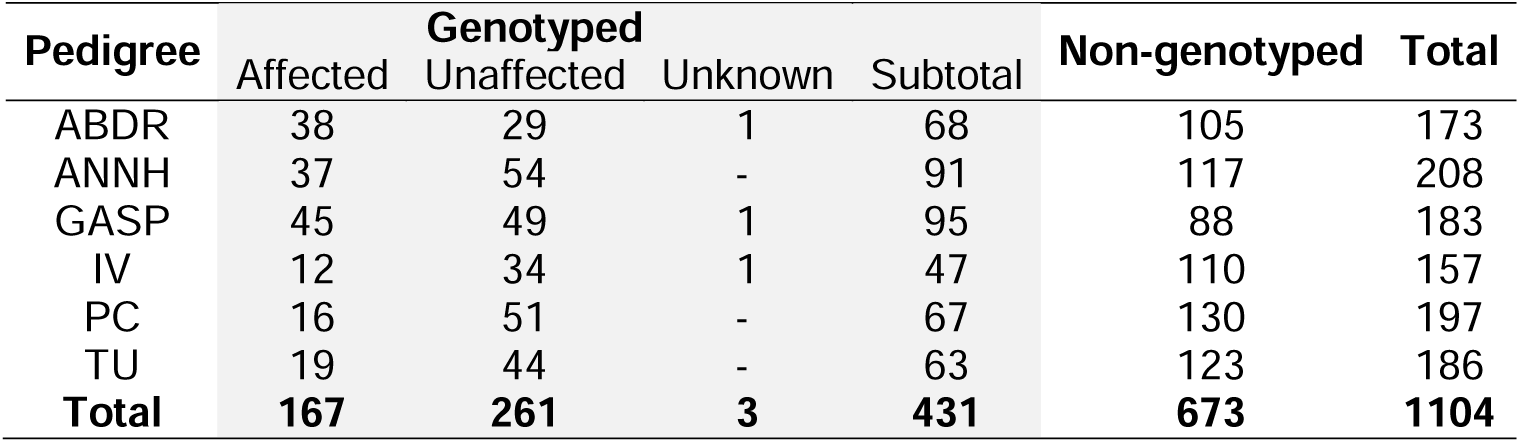
Distribution of samples across pedigrees. Total samples included by pedigree. Samples are separated into genotyped (affected, unaffected and unknown phenotype) and non-genotyped. Abobral (ABDR), André Lopes and Nhunguara (ANNH), Galvão and São Pedro (GASP), Ivaporunduva (IV), Pedro Cubas (PC) and Sapatu (TU).

| Pedigree | Genotyped |  |  |  | Non-genotyped | Total |
| --- | --- | --- | --- | --- | --- | --- |
|  | Affected | Unaffected | Unknown | Subtotal |  |  |
| ABDR | 38 | 29 | 1 | 68 | 105 | 173 |
| ANNH | 37 | 54 | - | 91 | 117 | 208 |
| GASP | 45 | 49 | 1 | 95 | 88 | 183 |
| IV | 12 | 34 | 1 | 47 | 110 | 157 |
| PC | 16 | 51 | - | 67 | 130 | 197 |
| TU | 19 | 44 | - | 63 | 123 | 186 |
| **Total** | **167** | **261** | **3** | **431** | **673** | **1104** |

**Table 2.** Study cohort characteristics. Comparison of cohort characteristics between affected and unaffected individuals across six pedigrees Abobral (ABDR), André Lopes and Nhunguara (ANNH), Galvão and São Pedro (GASP), Ivaporunduva (IV), Pedro Cubas (PC) and Sapatu (TU). The table presents the absolute number and percentage of males, mean age, systolic and diastolic blood pressure (BP) in mmHg, height in meters (m), weight in kilograms (kg), waist girth (cm), and hip girth (cm) for both affected and unaffected groups. The total number of individuals in each group is also provided for each pedigree. Values are shown as mean ± standard deviation. Pedigrees ABDR, GASP, and IV each include additionally one individual with an unknown phenotype.

|  | ABDR |  | ANNH |  | GASP |  | IV |  | PC |  | TU |  |
| --- | --- | --- | --- | --- | --- | --- | --- | --- | --- | --- | --- | --- |
|  | Affected | Unaffected | Affected | Unaffected | Affected | Unaffected | Affected | Unaffected | Affected | Unaffected | Affected | Unaffected |
| **Males** | 19 (50%) | 9 (31%) | 8 (21.6%) | 26 (48.1%) | 24 (53.3%) | 24 (49%) | 4 (33.3%) | 12 (35.3%) | 9 (56.2%) | 20 (39.2%) | 8 (42.1%) | 19 (43.2%) |
| **Age** | 49.7 ± 19.5 | 35.9 ± 15.9 | 57 ± 16.6 | 36.4 ± 16.7 | 54.3 ± 16.8 | 37 ± 13.5 | 64.3 ± 12.7 | 33.2 ± 14.1 | 60.6 ± 17 | 38.4 ± 14.6 | 53.8 ± 15.1 | 38.2 ± 17.2 |
| **Systolic BP** | 147.2 ± 25.9 | 121.6 ± 9.4 | 145.7 ± 23.2 | 112.1 ± 14.5 | 145.2 ± 21 | 120 ± 15.3 | 143.8 ± 18.6 | 115.9 ± 9.9 | 143.2 ± 15.4 | 128.7 ± 24.9 | 141.3 ± 21.8 | 112.5 ± 15.3 |
| **Diastolic BP** | 93.3 ± 13.4 | 75.1 ± 9.7 | 87.2 ± 11.5 | 75.9 ± 8.2 | 87.6 ± 14.4 | 75.1 ± 9.4 | 85.4 ± 11.6 | 73.5 ± 7.3 | 82.1 ± 11.4 | 80.9 ± 15.5 | 86.3 ± 6.1 | 73.3 ± 7.8 |
| **Height** | 1.6 ± 0.1 | 1.61 ± 0.1 | 1.54 ± 0.09 | 1.6 ± 0.12 | 1.58 ± 0.1 | 1.62 ± 0.09 | 1.57 ± 0.1 | 1.61 ± 0.08 | 1.6 ± 0.08 | 1.6 ± 0.09 | 1.6 ± 0.1 | 1.61 ± 0.08 |
| **Weight** | 63.3 ± 10.3 | 63.3 ± 10.7 | 58.5 ± 12.4 | 62.6 ± 12.8 | 64 ± 11.5 | 65.6 ± 12.3 | 65.7 ± 12.4 | 64.8 ± 11 | 64.3 ± 9.8 | 61.8 ± 12.9 | 69.8 ± 15.4 | 57.6 ± 9.5 |
| **Waist Girth** | 84.1 ± 9.5 | 83.5 ± 9.9 | 82.9 ± 10 | 80.2 ± 10 | 86.9 ± 9.9 | 83 ± 10.2 | 89.8 ± 10.5 | 82.8 ± 9.9 | 85.7 ± 5.5 | 82 ± 9.7 | 89.5 ± 8.1 | 76.3 ± 8.3 |
| **Hip Girth** | 94.3 ± 10.9 | 93.9 ± 11.8 | 93.4 ± 10 | 89.3 ± 10.9 | 94.7 ± 10.2 | 92.4 ± 12.7 | 97.7 ± 11.7 | 93.7 ± 14.3 | 90.6 ± 7.9 | 92.2 ± 10.5 | 97.5 ± 9.8 | 87.1 ± 8.6 |
| **Total** | **38 (55.9%)** | **29 (42.6%)** | **37 (40.7%)** | **54 (59.3%)** | **45 (47.4%)** | **49 (51.6%)** | **12 (25.5%)** | **34 (72.3%)** | **16 (23.9%)** | **51 (76.1%)** | **19 (30.2%)** | **44 (69.8%)** |

We filtered and trimmed the dataset (details in Data S1) using KING-Robust v.2.2.8^60^ and PLINK v.1.9/v.2.0^64^, a widely used tool for GWAS and population data analysis that efficiently handles large-scale data and performs quality control and kinship analysis. We excluded samples with a genotyping rate ≤ 95%, as well as SNPs with a genotyping rate ≤ 95%. Monomorphic SNPs and SNPs resulting in heterozygous haploid calls across all remaining individuals were removed. SNPs not adhering to Hardy-Weinberg equilibrium (*p*-value < 1×10^-3^) were also removed. SNP identification followed the dbSNP standard format (rsID), and their genetic locations (cM) were obtained through the Rutgers Combined Linkage-Physical Map v.3^65^. EH was considered a binary outcome, categorizing individuals as hypertensive (SBP ≥ 140 and/or DBP ≥ 90 mmHg) or normotensive (SBP < 140 and DBP < 90 mmHg). Individuals diagnosed and/or under medication for EH were classified as hypertensive.

We analyzed age differences between affected and unaffected individuals across pedigrees using Welch two sample t-tests, one-way and two-way ANOVA, and multiple regression analysis. These analyses were conducted using in-house R scripts, leveraging the R stats package v.3.6.2 (R Core Team, 2024) for statistical computations.

### LOCAL ANCESTRY ESTIMATION

We estimated local ancestry fractions (Figure 1, box F), which allowed us to determine individual- and pedigree-specific ancestries. The reference dataset comprised 189 samples from the 1000 Genomes Project^66^ and Stanford HGDP SNP Genotyping^67^ data: 63 European (CEU - Northern Europeans from Utah), 63 African (YRI - Yoruba in Ibadan, Nigeria), and 63 Native American (Colombia, Maya, and Pima populations) samples. Overlapping markers (145,467 SNPs) present in both reference (189 samples) and inference (431 samples) datasets were extracted and both datasets were merged and pruned for missingness (≤ 95% genotyping rate) using PLINK. Haplotypes were inferred using SHAPEIT2 v2.17,^68^ a tool for phasing genotype data and efficiently reconstructs haplotypes from large-scale datasets. RFMix v.1.5.4^69^, a tool for local ancestry inference, specializes in assigning ancestry to specific genomic segments in admixed populations was used. We used in-house scripts to estimate global ancestry fractions for each sample and pedigree ancestries by averaging the local ancestry calls across the entire genome (details in Data S2).

### PEDIGREE ANALYSIS

In our multipoint pedigree linkage analyses, we employed MORGAN suite, leveraging its versatility and robust capabilities rooted in the Markov Chain Monte Carlo (MCMC) approach, allowing for simultaneously handling numerous markers and individuals within pedigrees through a sampling methodology. ^61^

We implemented a redundant and semi-independent design to yield results that are both independent and comparable. From the complete inference marker set, we selected three distinct non-overlapping subsets (or subpanels) of markers (Figure 1, box E) employing specific selection criteria and parameters through PBAP (details in Data S3). The first subpanel, labeled the gold standard, included top-quality markers meeting specific criteria. The other two subpanels, while informative, might have slightly lower quality and were applied for validation of the finding. All three subpanels were crucial for discovery analyses. Parametric linkage analysis was performed using an autosomal-dominant model with a risk allele frequency of 0.01, an incomplete penetrance of 0.70 (for genotypes with 1 or 2 copies of the risk allele), and a phenocopy rate of 0.05. The penetrance was estimated by assessing affected and unaffected individuals within pedigrees. All subsequent steps were performed concurrently for each pedigree (details in Data S3.1).

We estimated allelic frequencies for each SNP marker (Figure 1, box G) using the specialized script ADMIXFRQ v.1.^53^ This involved the generation of unique pedigree-specific files organized by ancestry based on local ancestry calls and subpanel information.

To compute LOD scores, we employed the approach in MORGAN outlined as:

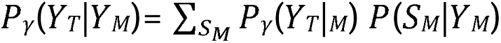

where *Y*_*M*_ denotes the genetic marker loci, *Y*_*T*_ the trait characteristics and *SM* denotes the meiosis indicators for all markers.^62^ The LOD score calculation was a two-step process. In the initial step (Figure 1, box H), we sampled inheritance vectors (IVs) for the gold standard subpanel using alternate SNP markers (2,500-3,000 SNPs) in a genome-wide “scan” analysis. Preliminary candidate regions were identified as those with a peak LOD score ≥ 1.

In the second step, we conducted a “dense” analysis (Figure 1, box J) restricted to the entire chromosomes containing each preliminary candidate region using all three subpanels and sampling of IVs using all available SNP markers. LOD scores were calculated again for this second step, defining Regions of Interest (ROIs) as those with a maximum LOD score ≥ 1.50 in at least one subpanel, with mandatory positive results for subpanel 1. ROI boundaries were determined based on LOD score ≥ 1 marker position (details in Data S3.3). Both steps (“scan” and “dense” analysis) were conducted separately for each pedigree.

To ensure the convergence of the sampling process, we performed diagnostic analysis of the MCMC runs (Figure 1, box I), evaluating run length, autocorrelation of LOD scores, and run stability using three different graphical tools (Figure S1). The default setup for MCMC runs was 100,000 Monte Carlo (MC) iterations, 40,000 burn-in iterations, 25 saved realizations, 1,000 identity-by-descent graphs and output scores saved at every 25 scored MC iterations (details in Data S3.4).

Results of this analysis were used to determine the appropriate running conditions. Once the correct setup for each pedigree was established, we repeated the genome-wide scan and dense analysis, which included sampling of IVs and calculation of LOD scores, to determine the final ROIs (Figure 1, box K).

### FINE-MAPPING STRATEGIES

Following pedigree analysis adjusted for admixture, we identified and fine-mapped 22 ROIs (Figure 1, box L-S). We further explored these results by analyzing the imputed SNP dataset using family-based association studies, individually evaluating each of the 22 ROIs while combining all pedigrees to increase statistical power. Additionally, we identified EH-related genes through *in-silico* investigation.

#### Family-Based Association Studies

Initially, we addressed population structure through principal components analysis (PCA) (Figure 1, box L), first employing MORGAN Checkped^61,62,70^ and PBAP Relationship Check^63^ algorithms for sample relatedness verification. Pairwise kinship coefficients (*Φ*) were calculated using KING-robust.^60^ The subsequent steps involved the iterative use of PC-AiR and PC-Relate functions conducted using the GENESIS (Genetic Estimation and Inference In Structured Samples) v3.19 R package.^57^ GENESIS provides tools for GWAS and offers advanced methods for handling relatedness and population structure in complex datasets. In the initial iteration, KING-robust estimates informed both kinship and ancestry divergence calculations, with resulting principal components (PCs) used to derive ancestry-adjusted kinship estimates, the 1^st^ genetic relationship matrix (GRM), via PC-Relate. To further refine PCs for ancestry, a secondary PC-AiR run utilized the 1^st^ GRM for kinship and KING-robust estimates for ancestry divergence, yielding new PCs. These new PCs informed a secondary PC-Relate run, culminating in a 2^nd^ GRM. Subsequently, we evaluated variation using the top 10 PCs, selected in accordance with the Kaiser criterion on the Kaiser-Guttman rule, which suggests retaining PCs with eigenvalues greater than 1, as these components explain more variance than a single original variable.^71^

We employed a comprehensive two-way independent imputation strategy (Figure 1, box N) for variants within each ROI identified through the family analysis (details in Data S4.2). We utilized linkage disequilibrium (LD) information to perform a population-based approach. This involved chromosome-wise dataset separation, phasing through SHAPEIT2+duoHMM method^72^, imputation using the Minimac4 v.4.0^73^ software and filtering (r^2^ ≥ 0.3) through BCFtools v1.15^74^. Minimac4 efficiently imputes genotypes from large datasets based on reference panels, while BCFtools provides utilities for processing and filtering variant call format (VCF) and binary call format (BCF) files. Concurrently a pedigree-based strategy incorporating IVs information was applied. This involved chromosome-wise dataset separation and imputation conducted using GIGI2 v.1^75^ a software tool designed for genetic imputation based on pedigree data.

To account for the complex correlation structure of the data, we included the 2^nd^ GRM as a random effect to fit the mixed models through fitNullModel function, conducted using the GENESIS R package^57^. Comprehensive testing included 10 ancestry principal components and all available covariates as fixed effects (details in Data S4.1). The final statistical model incorporated the top 8 PCs, along with sex, BMI (Body Mass Index), and age as significant variables. The next step was to fit the generalized linear mixed model (GLMM) (Figure 1, box M). Specifically for single-variant tests we employed a logistic mixed model expressed as:

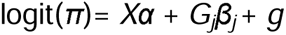

where *π* = *P*(*y* = 1 ∣ *X*,*G_j_*,*g)* represents the *Nx*1 column vector of probabilities of being affected for the *N* individuals conditional to covariates, allelic dosages; and random effects; *X* is the vector of covariates; and *α* is the vector of fixed covariate effects. We assume that 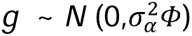 is a vector *g = (g_1,…,_ g_N_)* of random effects for the *N* subjects, where 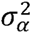 the additive genetic variance and Φ is the GRM; *G_j_* is a vector with the allelic dosages (0, 1, or 2 copies of the reference allele) or expected dose (in the case of imputed genotypes) at the locus *j*; and *β_j_* is its corresponding effect size. The null hypothesis of *β_j_* =0 was assessed using a multivariate score test^76^.

Finally, for each ROI, two independent FBAS tests were conducted (Figure 1, box O) using imputed variant datasets that are either (1) pedigree-based or (2) population-based. The dataset comprised all 431 samples from the combined 6 pedigrees. The single-variant association tests were conducted using the GENESIS R package^57^, implementing the adjusted GLMM to perform Score tests.

We performed multiple testing correction (Figure 1, box P) using the effective number of independent markers (*M_e_*) estimated using the Genetic Type I Error Calculator software v.0.1^77^, a software tool designed to assess the likelihood of false positives in GWAS. We also evaluated adequacy of the analysis modeling through evaluation of the genomic inflation factor (λ) by dividing the median of the chi-square statistics by the median of the chi-square distribution with 1 degree of freedom^78^. Analysis was carried out for each imputed variant dataset.

Suggestive and significant association variants underwent curation, assessment, and compilation (Figure 1, box Q) into a comprehensive database using a custom R script. Data were extracted in March 2023 from NIH dbSNP^79^, CADD^80^, NCBI PubMed^81^, Ensembl Variant Effect Predictor^82^, ClinVar^83^, Mutation Taster^84^, SIFT^85^, PolyPhen^86^, and VarSome^87^. Annotation included chromosome and physical positions (GRCh37/hg19), dbSNP rsID, associated genes, genomic alterations, variant consequences, exonic functions, and pathogenicity classifications. This procedure was executed independently for each ROI and for both imputed variant datasets.

Furthermore, we analyzed linkage disequilibrium (LD) (Figure 1, box R) patterns for variants within a 500,000 base pair window surrounding each statistically suggestive or significant associated variant. Utilizing the Ensembl REST API in conjunction with the ensemblQueryR tool v2.0^88^, an R package designed for efficient querying of the Ensembl database, we accessed and retrieved genomic data extracted during May 2024. We established thresholds of r^2^ ≥ 0.7 and D’ ≥ 0.9. This analysis incorporated data from the 1000 Genomes Project^66^ for multiple populations, including European (CEU: Utah Residents with Northern and Western European Ancestry), African (YRI: Yoruba in Ibadan, Nigeria), and American populations: CLM (Colombian in Medellin, Colombia), MXL (Mexican Ancestry in Los Angeles, California), PEL (Peruvian in Lima, Peru), and PUR (Puerto Rican in Puerto Rico). Additionally, we utilized the Ensembl REST API VEP to annotate and select LD patterns based on variant consequences.

#### Investigation of EH-related Genes

To elucidate the hypertension-related implications of genes identified within each ROI (Figure 1, box S), we first identified all genes mapped within ROIs using R package biomaRt v.2.6.0.1^89^. Then we obtained and annotated genes associated with “essential hypertension” or “high blood pressure” according to NCBI PubMed^81^, MedGen^90^, MalaCards^91^, ClinVar^83^, Ensembl BioMart^92^, and GWAS Catalog^93^. The data were extracted in March 2023. Finally, we matched both lists, systematically annotating based on physical position (base pair), cytogenetic band, summary, molecular function, related phenotype, gene ontology, genetic location (cM; GRCh37/hg19), expression patterns, and publication data. Additionally, to prioritize these genes (details in Data S5), we utilized VarElect v.5.21^94^, a tool that assesses the potential pathogenicity of genetic variants for prioritization.

#### Genetic Correlation Analysis

To validate our FBAS findings, we performed a genetic correlation analysis using summary statistics from our study and the publicly available GWAS Catalog dataset GCST90436066^95^, selected due to its extensive SNP coverage. The analysis was conducted using the GenomicSEM R package^96^, with LD scores calculated using LDSC scripts^97^ from the HapMap3 b36 YRI (Yoruba in Ibadan, Nigeria) population as the reference^98^. Given the high African ancestry in our dataset, the YRI population was selected as the reference for LD scores. Attempts to use LD scores from other HapMap3 populations resulted in negative heritability estimates, preventing accurate genetic correlation computation.

## RESULTS

Significant differences in mean ages were observed between affected and unaffected individuals (Figure 3) across all pedigrees (ABDR: 35.9 vs. 49.7 years, *p* = 2.112×10^-3^; ANNH: 36.4 vs. 57 years, *p* = 1.291×10^-7^; GASP: 37 vs. 54.3 years, *p* = 4.771×10^-7^; PC: 38.4 vs. 60.6 years, *p* = 1.026×10^-4^; IV: 33.2 vs. 64.3 years, *p* = 4.963×10^-7^; TU: 38.2 vs. 53.8 years, *p* = 8.866×10^-4^), as seen in Table S1. Overall, the combined data showed a mean age of 36.73 ± 15.4 years for unaffected individuals versus 55.11 ± 17.3 years for affected individuals (*p* = 6.855×10^-25^). One-way ANOVA was performed with 5 degrees of freedom for pedigrees and 161 for residuals, the sum of squares for pedigrees was 2783 and for residuals was 46617, resulting in mean squares of 556.6 and 289.5, respectively. Comparing the average age of affected individuals across pedigrees for both sexes, this analysis yielded an F value of 1.922 with a *p*-value of 0.093, indicating non-significant differences in average age across pedigrees. Two-way ANOVA highlighted a significant effect of affected status on average age (*p* < 0.001). However, neither sex nor the interaction between sex and affected status showed significant effects on average age, with *p*-values of 0.601 and 0.671 respectively. This suggests that while affected status significantly influences average age and sex, the interaction between sex and affected status does not have a significant impact. The multiple regression model showed that affected status significantly influenced average age (*p* < 0.001). The coefficients for sex (*p* = 0.197), pedigree (*p* = 0.551), BMI (*p* = 0.328), as well as for SBP (*p* = 0.278) and DBP (*p* = 0.440) are not statistically significant. The overall model’s performance is high (strong explanatory power) as indicated by the R-squared value of 0.761, suggesting that about 76.11% of the variability in average age can be explained by the predictors in the model. Additionally, the F-statistic is significant (*p* = 9.109×10^-5^), indicating that the model is significant in predicting average age.

**Figure 3.**
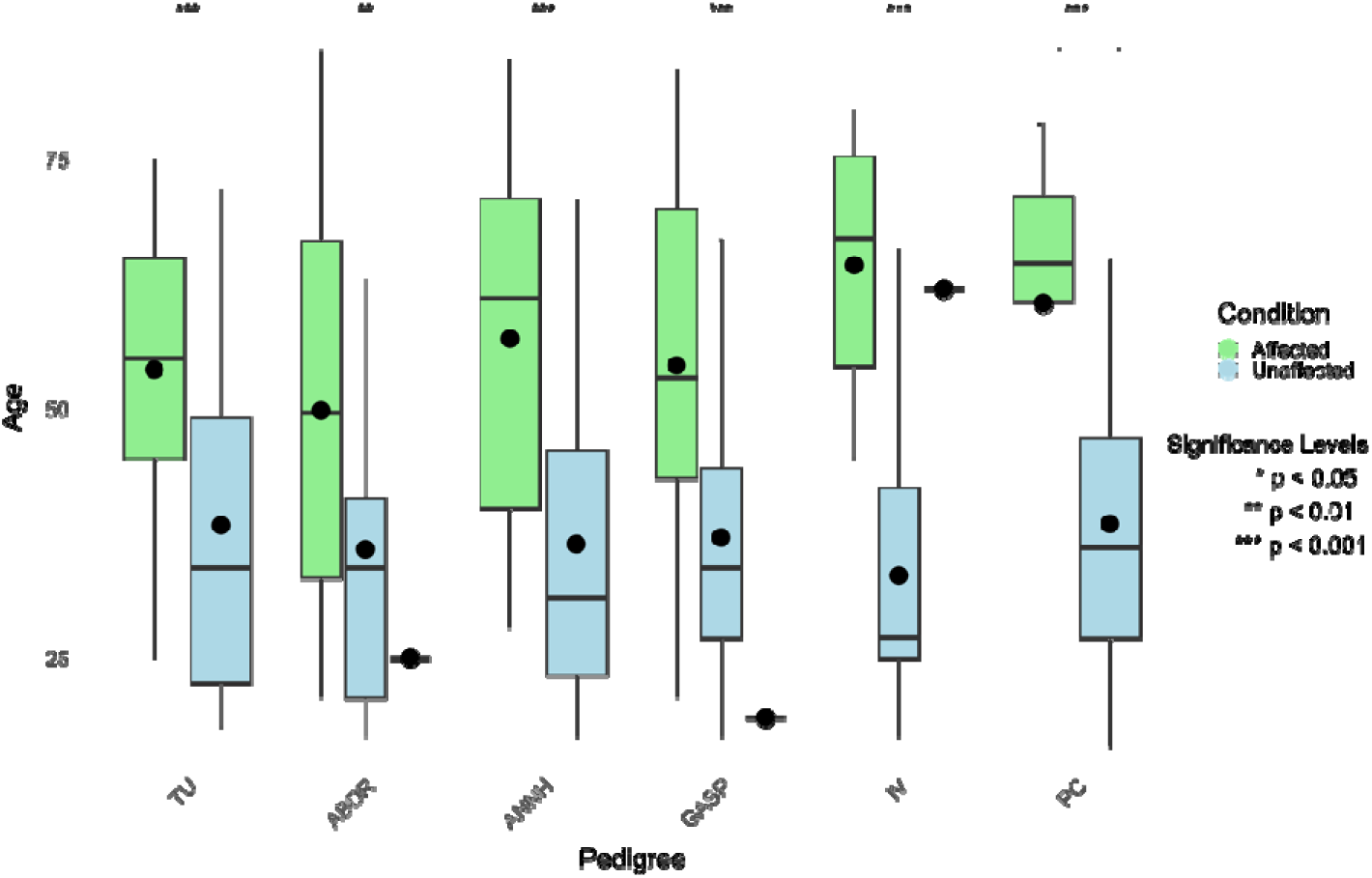
Distribution of ages for both affected and unaffected individuals across different pedigrees. The box plots represent the interquartile range (IQR) with the median age indicated by the line within each box. The small black dots indicate the mean ages for each group, and the error bars represent the standard deviation around these means. Affected individuals are shown in green, and unaffected individuals are shown in blue. Pedigrees Abobral (ABDR), Galvão and São Pedro (GASP), and Ivaporunduva (IV) each include additionally one non-represented individual with an unknown phenotype.

We calculated ancestry proportions for individual admixture segments (local ancestry) using data from 145,467 SNPs considering all the 431 samples (Figure 4A) and individually per pedigree (Figure 4B). The three top principal components (PCs), PC1, PC2 and PC3, explain 12.42%, 6.17%, and 1.68% of the variance, respectively (Figure 5A), allowing to visualize the genetic distance between the inference and the reference datasets (Figure 5B). All six quilombo remnants pedigrees studied have a high degree of admixture. Among all 431 individuals the estimates are 47.4%, 36.3%, and 16.1%, respectively, for African, European, and Native American ancestries (Table 3). The pedigrees from ABDR and PC had the highest (51.5%) estimates of African ancestral contribution. In comparison, TU had the highest (50.8%) estimate of European ancestral contribution, and ABDR had the highest estimate (16.9%) of Native American ancestral contribution. For comparison, the ancestry proportions for the general Brazilian population were estimated as 19.6%, 68.1%, and 11.6% for African, European, and Native American ancestral contribution^50^, respectively, with vivid contrasts among each Brazilian region (Table S2-3).

**Figure 4.**
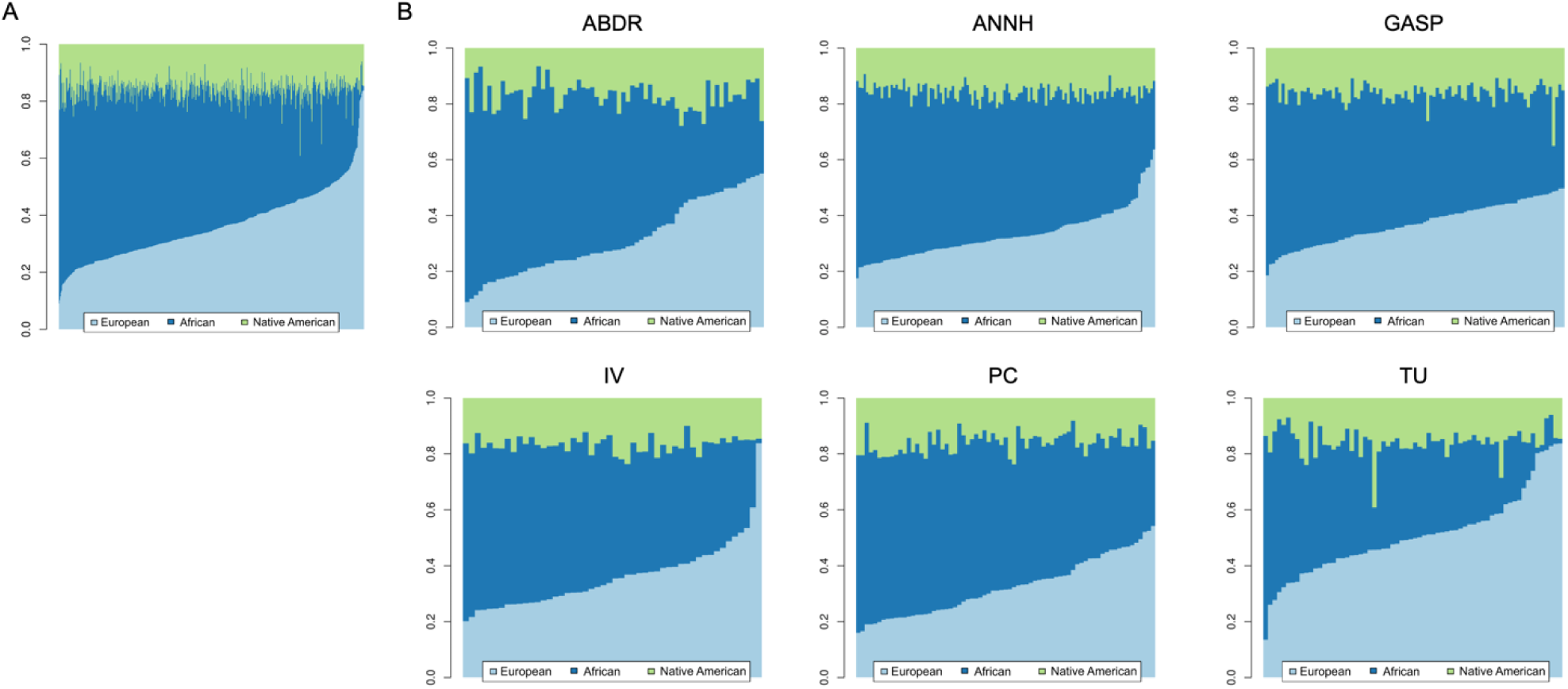
Graphic representation of global ancestry estimates. A) Ancestry estimates for the entire dataset (n = 431); B) Ancestry estimates stratified by pedigree. Colors represent ancestral contributions: green (Native American), dark blue (African), and light blue (European). Pedigrees include Abobral (ABDR), André Lopes and Nhunguara (ANNH), Galvão and São Pedro (GASP), Ivaporunduva (IV), Pedro Cubas (PC), and Sapatu (TU).

**Figure 5.**
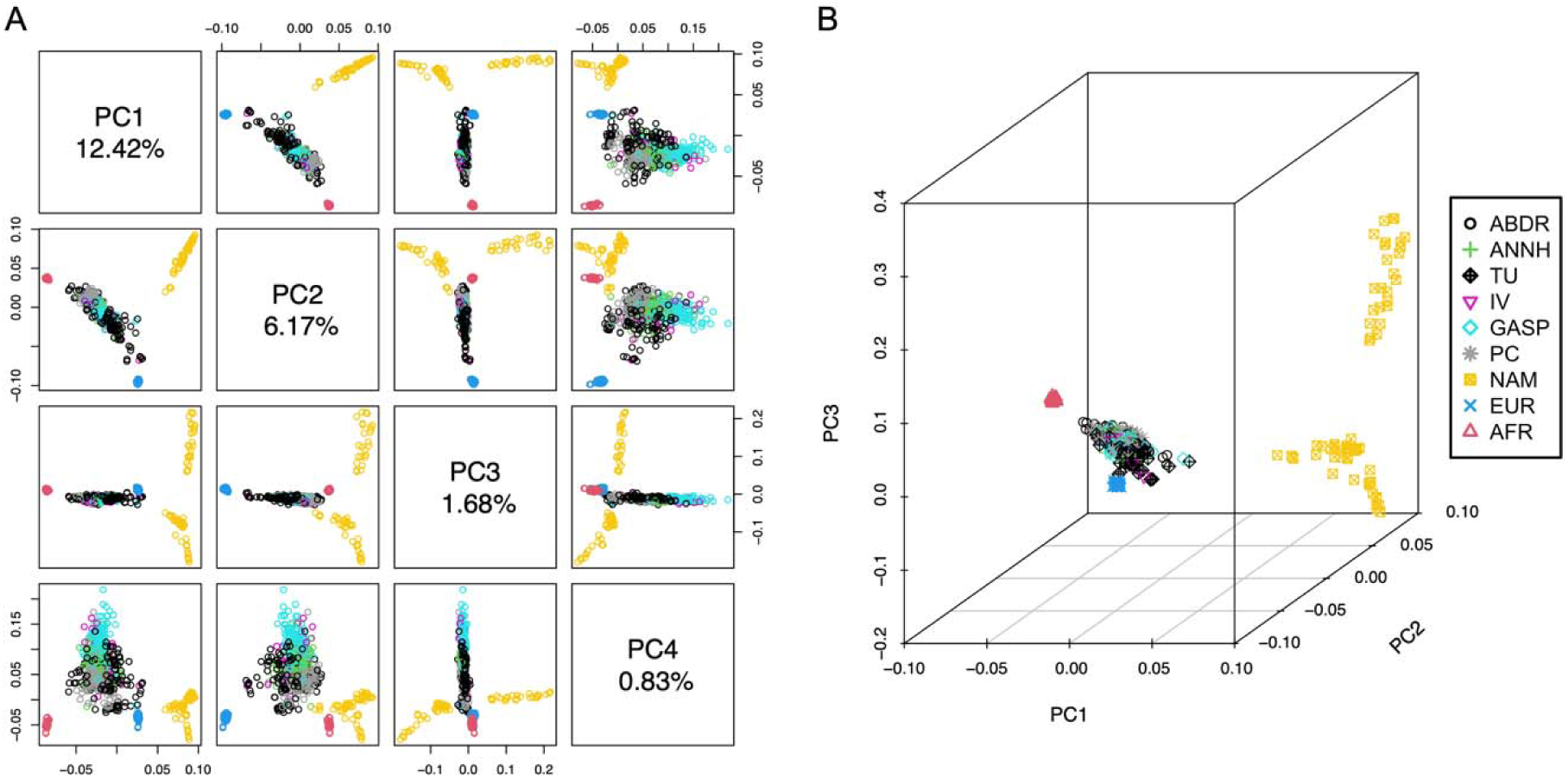
Principal Component Analysis (PCA) of Genetic Distance. A) PCA results for the first four principal components (PCs); B) Three-dimensional PCA representation. Reference samples from European (EUR), African (AFR), and Native American (NAM) populations are depicted in blue, red, and yellow, respectively. Pedigrees include Abobral (ABDR), André Lopes and Nhunguara (ANNH), Galvão and São Pedro (GASP), Ivaporunduva (IV), Pedro Cubas (PC), and Sapatu (TU).

**Table 3.** Ancestry Proportions. The ancestry proportions (%) are presented for each pedigree, detailing the contributions of African, European, and Native American ancestries. The corresponding sample sizes are included for reference. The total dataset is summarized under “Total.” Pedigrees are identified as Abobral (ABDR), André Lopes and Nhunguara (ANNH), Galvão and São Pedro (GASP), Ivaporunduva (IV), Pedro Cubas (PC), and Sapatu (TU).

| Pedigrees | Sample size | Ancestry Proportions |  |  |
| --- | --- | --- | --- | --- |
|  |  | African | European | Native American |
| ABDR | 68 | 51.5% | 31.4% | 16.9% |
| GASP | 95 | 46.8% | 36.9% | 16.2% |
| ANNH | 91 | 50.8% | 33.2% | 15.8% |
| IV | 47 | 47.3% | 35.9% | 16.7% |
| TU | 63 | 33.4% | 50.8% | 15.7% |
| PC | 67 | 51.5% | 32.4% | 16.0% |
| **Total** | **431** | **47.4%** | **36.3%** | **16.1%** |

The MCMC runs diagnosis were conducted, within each pedigree, on the smallest chromosome exhibiting a positive LOD score (Figure S2-7). Through pedigree analysis, we identified 30 ROIs (Table S4) of which 8 were discarded due to a lack of corroboration between the three subpanels of SNPs. The remaining 22 ROIs contain 2363 genes (Table S5) (example in Figure 6A).

**Figure 6.**
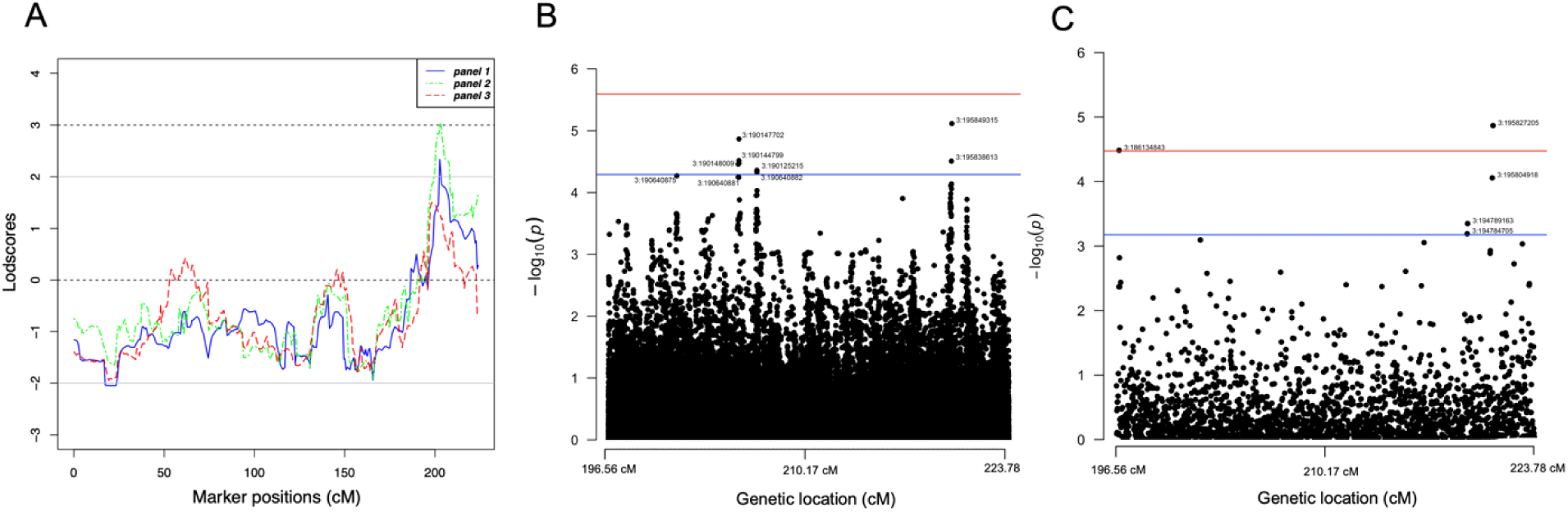
Illustration of the results for ROI5. A) Dense linkage analysis across the three subpanels; B) Association study using population-based imputed data; C) Association study using pedigree-based imputed data. Black dots represent SNPs (labeled by chromosome and physical position), blue lines indicate the suggestive *p*-value threshold (-log10(p)) and the red lines to the significant *p*-value threshold (-log10(p)).

We utilized family-based association studies (Figure 6B-C) to analyze a total of 1,612,754 SNPs, derived from both pedigree- and population-based imputation strategies, across the 22 ROIs. The summary statistics for the extended SNP set are publicly accessible in the GWAS Catalog (GCST90454187). From this analysis, we identified 117 variants (Table S6) with suggestive or significant associations with EH.

Furthermore, our investigation of EH-related genes, utilizing publicly available databases and supported by the literature, narrowed down the initial set of 2363 genes to 60 promising candidate genes (Table 4) located within the 22 ROIs.

**Table 4.** Key Genes Identified Across Analysis Strategies. Main genes identified from the three analysis strategies—linkage analysis, EH-related gene investigation (EH), and association studies (AS)—for each region of interest (ROI).

| Gene | Cytoband | ROI | EH | AS |
| --- | --- | --- | --- | --- |
| *ALPK2* | 18q21.31-q21.32 | 20 | • | • |
| *EDARADD* | 1q42.3 | 3 | • | • |
| *KCNT1* | 9q34.3 | 11 | • | • |
| *LPP* | 3q27.3-q28 | 5 | • | • |
| *MFN2* | 1p36.22 | 2 | • | • |
| *MTR* | 1q43 | 3 | • | • |
| *P2RX1* | 17p13.2 | 18 | • | • |
| *PHGDH* | 1p12 | 1 | • | • |
| *RPA1* | 17p13.3 | 18 | • | • |
| *RYR2* | 1q43 | 3 | • | • |
| *S100A10* | 1q21.3 | 1 | • | • |
| *SERTAD2* | 2p14 | 4 | • | • |
| *TENM4* | 11q14.1 | 13 | • | • |
| *ZZEF1* | 17p13.2 | 18 | • | • |
| *ABCC9* | 12p12.1 | 14 | • |  |
| *ACE* | 17q23.3 | 19 | • |  |
| *ADIPOQ* | 3q27.3 | 5 | • |  |
| *APOE* | 19q13.32 | 21 | • |  |
| *ARHGEF17* | 11q13.4 | 13 | • |  |
| *BORCS7* | 10q24.32 | 12 | • |  |
| *CASP3* | 4q35.1 | 7 | • |  |
| *CDH13* | 16q23.3 | 17 | • |  |
| *CNNM2* | 10q24.32 | 12 | • |  |
| *CORIN* | 4p12 | 6 | • |  |
| *CYGB* | 17q25.1 | 19 | • |  |
| *CYP17A1* | 10q24.32 | 12 | • |  |
| *CYP2C19* | 10q23.33 | 12 | • |  |
| *CYP2C9* | 10q23.33 | 12 | • |  |
| *EDN1* | 6p24.1 | 9 | • |  |
| *HTR2A* | 13q14.2 | 16 | • |  |
| *KDR* | 4p12 | 6 | • |  |
| *KIT* | 4p12 | 6 | • |  |
| *KLKB1* | 4q35.2 | 7 | • |  |
| *KNG1* | 3q27.3 | 5 | • |  |
| *LPL* | 8p21.3 | 10 | • |  |
| *MC4R* | 18q21.32 | 20 | • |  |
| *MEIS1* | 2p14 | 4 | • |  |
| *NT5C2* | 10q24.32-q24.33 | 12 | • |  |
| *PDE3A* | 12p12.2 | 14 | • |  |
| *PDE4D* | 5q11.2-q12.1 | 8 | • |  |
| *PHACTR1* | 6p24.1 | 9 | • |  |
| *PLCE1* | 10q23.33 | 12 | • |  |
| *PRKCA* | 17q24.2 | 19 | • |  |
| *RBM47* | 4p14 | 6 | • |  |
| *RBP4* | 10q23.33 | 12 | • |  |
| *SGCZ* | 8p22 | 10 | • |  |
| *SLC1A4* | 2p14 | 4 | • |  |
| *SLC39A14* | 8p21.3 | 10 | • |  |
| *SOCS3* | 17q25.3 | 19 | • |  |
| *SST* | 3q27.3 | 5 | • |  |
| *TBX2* | 17q23.2 | 19 | • |  |
| *TGFB1* | 19q13.2 | 21 | • |  |
| *TIMP2* | 17q25.3 | 19 | • |  |
| *TLR3* | 4q35.1 | 7 | • |  |
| *TMOD4* | 1q21.3 | 1 | • |  |
| *TNFRSF1B* | 1p36.22 | 2 | • |  |
| *UCP2* | 11q13.4 | 13 | • |  |
| *VEGFC* | 4q34.3 | 7 | • |  |
| *WBP1L* | 10q24.32 | 12 | • |  |
| *ZDHHC2* | 8p22 | 10 | • |  |

Among the 22 remaining ROIs, 20 of them were single-supported by the investigation of EH-related genes as a fine-mapping strategy, and 17 of them were additionally supported by family-based association studies as a fine mapping strategy (double-supported). Highlighting the common ground between the double supported results (by both the fine-mapping strategies), 14 genes (highlighted in Table 4) were identified within the mapped regions with compelling evidence of association with the phenotype. These genes include *PHGDH* and *S100A10* (ROI1); *MFN2* (ROI2); *RYR2*, *EDARADD*, and *MTR* (ROI3); *SERTAD2* (ROI4); *LPP* (ROI5); *KCNT1* (ROI11); *TENM4* (ROI13); *P2RX1*, *ZZEF1*, and *RPA1* (ROI18); and *ALPK2* (ROI20). All these genes were broadly described in the supplementary section (SS7: Double-supported EH genes). These genes harbor 29 SNPs (highlighted in Table S6) implicated by our family-based association studies.

From our analysis, the estimated genetic correlation between our study and GCST90436066 was 0.427, with a genetic covariance of 0.0105 (*p* = 0.743). Heritability estimates for GCST90436066 showed a total observed scale heritability (h²) of 0.0023 (*Z* = 3.15, *p* = 7×10^-4^), while for our study, h² was 0.2623 (Z = 0.271), reflecting a significant difference between the datasets. These results are likely influenced by the fact that GCST90436066 represents a European cohort (there is no available African or admixed EH summary statistics in public repositories such as the GWAS Catalog), in contrast to the admixed nature of our dataset, which contains a high proportion of African ancestry. Despite these limitations, the moderate genetic correlation observed provides some level of validation, indicating some level of shared genetic architecture between the two cohorts for this trait.

## DISCUSSION

Despite significant financial investment and the substantial number of samples analyzed in GWLA and GWAS, much of the heritability of complex phenotypes such as EH remains elusive. Moreover, due to the underrepresentation of African and Native American populations in global genetic studies, much of what is currently known about complex phenotypes may not be fully applicable to admixed populations, such as the descendants of quilombo populations and, more broadly, the Afro-descendant Brazilians. By studying EH in semi-isolated quilombo remnants, we reduce biases from population and clinical-biological heterogeneity, improve the signal-to-noise ratio (and consequently the statistical power), and increase the representation of admixed populations (like the Brazilian population) in genomic studies.

In our complementary approach, we combined two main strategies, i.e., linkage analysis and family-based association studies. Together, these strategies minimize the need for multiple testing, reduce population stratification, and leverage chromosomal location information provided by meiotic events. This combined approach resulted in 22 filtered ROIs with 60 single-supported genes (harboring 117 suggestive/significant associated variants) by the EH-genes investigation, and 14 double-supported genes by both the fine-mapping (investigation of EH-genes and FBAS) harboring 29 suggestive/significant associated variants.

The 14 double-supported genes imply EH through their roles in vascular, cardiac, and metabolic biological pathways. For instance, *LPP* harbors BP associated SNPs^99^ and plays a role in maintaining vascular smooth muscle cell contractility^100^, which helps prevent hypertension-induced arterial remodeling^101^. Similarly, *P2RX1*, identified in both African ancestry populations and our Brazilian Afro-descendant cohort, plays a key role in vasoconstriction in small arteries^102^ and renal autoregulation, both of which are critical for maintaining blood pressure homeostasis^103^. Apparently, this gene controls glomerular hemodynamics in ANG II hypertension^104^. *RYR2* harbors several BP and EH-associated SNPs^105–107^ and is key to calcium handling in cardiac muscle^102^, directly affecting heart function and blood pressure regulation, while *MFN2* is involved in mitochondrial function^102^, with mutations linked to vascular smooth muscle proliferation, which may contribute to hypertension^108–110^. Collectively, these genes are involved in mechanisms affecting vascular tone, muscle contraction, and energy metabolism, all central to blood pressure regulation.

Several genes related to oxidative stress, endothelial dysfunction, and nitric oxide (NO) regulation, were associated to EH in this study and supported by the literature. *RPA1* influences the repression of endothelial nitric oxide synthase (eNOS)^111^, thus reducing NO levels and leading to endothelial dysfunction^112^, a key player in hypertension. *EDARADD* has been linked to pulse pressure regulation^102,113^, while *S100A10* is involved in cellular processes such as inflammation^114–116^, which can contribute to vascular dysfunction and elevated blood pressure. *MTR*, responsible for methionine biosynthesis^102^, has been associated with diastolic blood pressure^117^, indicating its possible role in homocysteine metabolism and cardiovascular risk in EH patients^118^, and has been suggested as a genetic marker to assess the action of antihypertensive drugs^119^. We have identified a blood pressure -related SNP in *MTR* (rs1805087) in linkage disequilibrium (r^2^ ≥ 0.97) with the suggestive SNP rs10925261 (*p*-value = 2.5×10^-3^) identified in this study. This set of genes collectively suggests that endothelial health, inflammation, and metabolic regulation converge on pathways influencing blood pressure.

Lastly, genes such *as ZZEF1*^120,121^, *PHGDH*^122,123^, and *TENM4*^124,125^ were associated with blood pressure regulation via several genetic polymorphisms. *PHGDH* has been linked to blood pressure through epigenetic regulation, with methylation changes influencing systolic and diastolic pressure^123^. *KCNT1* may influence vascular tone indirectly although it is mainly involved in potassium channel regulation^126,127^. Although *SERTAD2*^110^ and *ALPK2*^128^ present BP regulation-associated polymorphisms, they have less direct evidence connecting them to EH but their involvement in cellular signaling and protein interactions suggest possible contributions to blood pressure regulation.

Note that the SNPs linked to the EH phenotype identified in this study were genotyped using genomic arrays and are therefore common variants. Many of these variants are situated in non-coding or intergenic regions and may or may not have recognized functional or regulatory effects. Although these SNPs are not expected to affect the phenotype directly, several of these variants are in LD with variants of more significant impact. Notably, among these, some may be rare and remain ungenotyped. Specifically, we identified unique tag SNPs: 77 non-coding SNPs (3’ UTR, 5’ UTR, or TF binding), 196 regulatory, and 15 missense SNPs (Table S7).

Recent GWAS studies have highlighted significant findings in African-derived populations, revealing genetic variants associated with blood pressure traits that differ from those identified in European populations. For example, a large meta-analysis involving 80,950 individuals of African ancestry identified 10 variants for SBP and 9 for DBP, including a novel variant, rs562545 in the *MOBP* gene, associated with DBP^129^. The AWI-Gen study, which focused on sub-Saharan African populations, identified two genome-wide significant signals for blood pressure traits: one near *P2RY1* for SBP and another near *LINC01256* for pulse pressure^130^. These findings, absent in European ancestry cohorts, demonstrate the critical need for African-derived specific studies. Similarly, large-scale GWAS efforts, such as those utilizing the UK Biobank, discovered that only a fraction of loci identified in European populations replicate in African-derived populations, demonstrating that ancestry-specific genetic variants play a crucial role in hypertension susceptibility^129,130^. African-derived populations often exhibit unique genetic associations due to differences in genetic architecture. For example, variants in genes such as *CACNA1D* and *KCNK3* that have been found to significantly contribute to blood pressure regulation in African populations are less prominent in other populations.

Admixture mapping has provided valuable insights into loci with potential effects on BP and EH, identifying genomic regions particularly relevant in populations with African ancestry. For instance, *NPR3*, associated with blood pressure regulation in both African and European populations, was uncovered through admixture mapping in African American^131^. Additional regions such as 1q21.2–21.3, 4p15.1, 19q12, and 20p13 have demonstrated significant associations with DBP (*p*-values ≤ 2.07×10^-4^), while regions 1q21.2–21.3 and 19q12 were also associated (*p*-values ≤ 5.32×10^-4^) with mean arterial pressure^132^.

Several regions identified in the literature have been corroborated by findings from our study. The 3q27.3 region (ROI 5), containing the *AGT* gene, is strongly linked to EH, especially in African ancestry populations^133^. Likewise, the 17q23.2 region (ROI 19) has been implicated in blood pressure traits, particularly through its role in regulating the RAAS^134^. Furthermore, 6p24.1 (ROI 9) has shown previous associations with blood pressure regulation^133^. Linkage studies have also implicated 1q43 (ROI 3) in essential hypertension^135^, while 8p23.1 (ROI 10) has been recognized for its relevance in blood pressure regulation, particularly in populations of African descent^134^.

These findings underscore the value of including diverse populations in genetic studies to uncover novel loci and better understand the genetic basis of complex traits like hypertension. In this study, our approach identified 22 ROIs, with 14 relevant genes, from which we highlight *PHGDH*, *S100A10*, and *RYR2* implicated in hypertension susceptibility. The overlap between findings in African ancestry populations and those in Brazilian Afro-descendants, such as *P2RX1* highlighted in our stud*y*, further supports the role of GWAS and admixture mapping in identifying population-specific loci relevant to hypertension risk.

To prioritize our results, we developed a comprehensive score based on the weights assigned to each strategy (Table S8), including linkage analysis, EH genes investigation, and association studies. The scores allowed to classify the ROIs into three tiers based on their priority level (Table 5): high (top 20% of the ROIs), intermediate (30% of the ROIs), and low (50% of the ROIs).

**Table 5.** Ranked Score-Weighted Regions of Interest (ROIs). The ROIs are classified into three tiers according to their priority level: the top 20% are labeled as high priority (dark gray), the medium 30% as intermediate priority (medium gray), and the bottom 50% as low priority (light gray).

| ROI | Cytoband | ROI size (Mb) | EH Genes | Linkage Analysis |  | Assoc. Studies Sugg./Sig. | Gene Relevance | TOTAL |
| --- | --- | --- | --- | --- | --- | --- | --- | --- |
|  |  |  |  | Peak LOD Score | Subpanels consensus |  |  |  |
| 5 | 3q27.3-q29 | 11.4 Mb | Yes \| 4 genes (+1p) | 3.036 (+6p) | 3 (+3p) | Sig. (+3p) | Yes (+5p) | 18 |
| 12 | 10q23.33-q25.1 | 11.2 Mb | Yes \| 9 genes (+1p) | 2.795 (+5p) | 3 (+3p) | Sig. (+3p) | Yes (+5p) | 17 |
| 13 | 11q13.4-q14.1 | 7.6 Mb | Yes \| 3 genes (+1p) | 2.414 (+4p) | 3 (+3p) | Sugg. (+1p) | Yes (+5p) | 14 |
| 19 | 17q23.2-25.3 | 19 Mb | Yes \| 9 genes (+1p) | 2.138 (+3p) | 3 (+3p) | Sugg. (+2p) | Yes (+5p) | 14 |
| 3 | 1q43 | 1.3 Mb | Yes \| 3 genes (+1p) | 2.161 (+3p) | 2 (+2p) | Sugg. (+2p) | Yes (+5p) | 13 |
| 9 | 6p24.1-p22.3 | 8 Mb | Yes \| 2 genes (+1p) | 2.621 (+5p) | 2 (+2p) | Sugg. (+2p) | Yes (+3p) | 13 |
| 10 | 8p23.1-p21.3 | 9.8 Mb | Yes \| 4 genes (+1p) | 2.658 (+5p) | 3 (+3p) | Sugg. (+1p) | Yes (+3p) | 13 |
| 7 | 4q32.3-q35.2 | 20.6 Mb | Yes \| 4 genes (+1p) | 2.322 (+4p) | 2 (+2p) | Sugg. (+2p) | Yes (+3p) | 12 |
| 21 | 19q13.12-13.32 | 10 Mb | Yes \| 2 genes (+1p) | 2.503 (+4p) | 3 (+3p) | Sugg. (+1p) | Yes (+3p) | 12 |
| 14 | 12p12.3-p11.23 | 11.1 Mb | Yes \| 2 genes (+1p) | 2.662 (+5p) | 2 (+2p) | Sugg. (+1p) | Yes (+2p) | 11 |
| 8 | 5q12.1-q13.2 | 11.1 Mb | Yes \| 1 gene (+1p) | 2.117 (+3p) | 3 (+3p) | Sugg. (+2p) | Yes (+2p) | 11 |
| 18 | 17p13.3-p13.2 | 4.3 Mb | Yes \| 3 genes (+1p) | 1.771 (+2p) | 2 (+2p) | Sugg. (+1p) | Yes (+4p) | 10 |
| 2 | 1p36.21-p36.22 | 3.2 Mb | Yes \| 2 genes (+1p) | 1.824 (+2p) | 3 (+3p) | Sugg. (+1p) | Yes (+3p) | 10 |
| 16 | 13q14.13-q21.33 | 25.6 Mb | Yes \| 1 gene (+1p) | 2.554 (+4p) | 3 (+3p) | No (0p) | Yes (+2p) | 10 |
| 1 | 1p12-q21.3 | 32.5 Mb | Yes \| 3 genes (+1p) | 1.503 (+1p) | 2 (+2p) | Sugg. (+1p) | Yes (+4p) | 9 |
| 4 | 2p14 | 3.4 Mb | Yes \| 3 genes (+1p) | 1.906 (+2p) | 2 (+2p) | Sugg. (+1p) | Yes (+3p) | 9 |
| 20 | 18q21.32 | 1.7 Mb | Yes \| 2 genes (+1p) | 1.608 (+1p) | 3 (+3p) | Sugg. (+1p) | Yes (+3p) | 9 |
| 6 | 4p15.1-q12 | 26.5 Mb | Yes \| 4 genes (+1p) | 1.938 (+2p) | 3 (+3p) | No (0p) | Yes (+3p) | 9 |
| 11 | 9q34.3 | 0.8 Mb | Yes \| 1 gene (+1p) | 1.418 (+1p) | 3 (+3p) | Sugg. (+1p) | Yes (+2p) | 8 |
| 17 | 16q23.3-q24.1 | 4.5 Mb | Yes \| 1 gene (+1p) | 2.175 (+3p) | 2 (+2p) | No (0p) | Yes (+2p) | 8 |
| 15 | 12q24.32-q24.33 | 4.6 Mb | No (0p) | 2.087 (+3p) | 2 (+2p) | Sugg. (+1p) | Yes (+1p) | 7 |
| 22 | 19q13.41-13.42 | 3.2 Mb | No (0p) | 1.860 (+2p) | 3 (+3p) | Sugg. (+1p) | Yes (+1p) | 7 |

As presented, our study has some limitations. One such limitation is that the investigation of EH-related genes, as part of the fine-mapping strategy, was focused on genes previously supported by literature as being related to blood pressure regulation. Consequently, it is possible that genes that have never been implicated in EH may contain rare or novel variants relevant to the origin of hypertension in quilombo remnant populations.

On the other hand, the strength of this study comes from the improvement and application of a unique multi-level computational approach that combined mapping strategies to deal with large family data, which provided reliable results. By using genome-wide linkage analyses based on MCMC methods and adjusted for admixture, association studies as the primary fine-mapping strategy, and limiting analyses to candidate genomic regions, this study took advantage of meiotic information provided by pedigrees while simultaneously reducing the need for multiple tests and avoiding population stratification. Therefore, the ROIs identified in this study are credible and provide valuable insights into the genetic basis of essential hypertension in the quilombo remnant populations.

Conducting analyses by merging all six pedigrees into only one would be a formidable challenge, and probably not feasible. However, the prospect of replicating these analyses using alternative computational packages is exciting and not an impossible task. Our study has demonstrated that blood pressure and hypertension in the quilombo remnant populations are likely influenced by multiple genes, in a polygenic or oligogenic mechanism of inheritance. We have identified several loci across different chromosomes that contain genes and variants involved in the development of hypertension. Additionally, we have identified genomic regions of interest not previously associated with EH and will therefore be important targets for future research.

To overcome the limitations, future steps of this investigation will involve the use of Whole Genome Sequencing (WGS) and Whole Exome Sequencing (WES) from this dataset. WGS and WES will enable the investigation of coding and non-coding variants within all ROIs, with a focus on rare variants that may have a higher impact on gene functioning. To optimize this process, the prioritization of ROIs as performed in Table 5 is essential, since high and intermediate ROIs will be addressed first when filtering variants detected after WES and WGS, and the low-priority ROIs afterwards.

Furthermore, our study has the potential to shed light on important genomic regions, genes, and variants that are specific to African-derived populations. We have provided insights into the genetic factors that contribute to hypertension in a group that has been often underrepresented in genetic studies and databases.

## Data Availability

All data produced in the present study are available upon reasonable request to the authors.

https://livemarshall-my.sharepoint.com/:b:/g/personal/magalhaesbor_marshall_edu/EfnaW0V_-aJPjyxSiTESfpkBXlK3cfPUBnFGL2KTiNLOQg?e=fbhHuz

## ACKNOWLEDGMENTS

Authors would like to thank the support of Dr. Diogo Meyer (IB/USP - Brazil), Dr. Julia M. Pavan Soler (IME/USP - Brazil), Dr. Suely Ruiz Giolo (UFPR - Brazil), Dr. Paulo Otto (IB/USP - Brazil) and Dr. Alexandre da Costa Pereira (INCOR - Brazil) for valuable suggestions. The authors also thank Dr. Christian Kubisch (University Medical Center Hamburg / Eppendorf - Germany) for many ideas which stimulated the development of this project.

## SUPPLEMENTAL MATERIAL

Data S1-S6

Tables S1-S8

Figures S1-S7

References 135-154

## NON-STANDARD ABBREVIATIONS AND ACRONYMS

ABDR: Abobral (quilombo population)
*AGT*: angiotensinogen
*ALPK2*: alpha kinase 2
AN: André Lopes (quilombo population)
ANG II: angiotensin II
ANP: atrial natriuretic peptide
*ARMC*: armadillo repeat containing 5
BMI: body mass index
BP: arterial blood pressure
*CACNA1D*: calcium voltage-gated channel subunit alpha1 d
CADD: combined annotation dependent depletion
CEU: Northern Europeans from Utah
CLM: Colombian in Medellin, Colombia
CVDs: cardiovascular diseases
*CYP11B2*: cytochrome p450 family 11 subfamily b member 2
DBP: diastolic blood pressure
*EDARADD*: edar associated via death domain
EH: essential hypertension
eNOS: endothelial nitric oxide synthase
GA: Galvão (quilombo population)
GLMM: generalized linear mixed model
*GRK4*: g protein-coupled receptor kinase 4
GRM: genetic relationship matrix
HGDP: human genome diversity project
IV: Ivaporunduva (quilombo population)
IVs: inheritance vectors
*KCNK3*: potassium two pore domain channel subfamily k member 3
*KCNT1*: potassium sodium-activated channel subfamily t member 1
KKS: kallikrein-kinin system
LD: linkage disequilibrium
*LINC01256*: long intergenic non-protein coding rna 1256
LOD: logarithm of the odds
*LPP*: lim domain containing preferred translocation partner in lipoma
MCMC: markov chain monte carlo
*MFN2*: mitofusin 2
MLX: Mexican Ancestry in Los Angeles, California
*MOBP*: myelin associated oligodendrocyte basic protein
*MTR*: 5-methyltetrahydrofolate-homocysteine methyltransferase
NH: Nhunguara (quilombo population)
NO: nitric oxide
*NOS3*: nitric oxide synthase 3
*NPR3*: natriuretic peptide receptor 3
*P2RX1*: purinergic receptor p2x 1
*P2RY1*: purinergic receptor p2y1
PC: Pedro Cubas (quilombo population)
PCA: principal components analysis
PCs: principal components
PEL: Peruvian in Lima, Peru
*PHGDH*: phosphoglycerate dehydrogenase
PHOX2: paired like homeobox 2
PUR: Puerto Rican in Puerto Rico
RAAS: renin-angiotensin-aldosterone system
ROIs: regions of interest
*RPA1*: replication protein a1
*RYR2*: ryanodine receptor 2
*S100A10*: s100 calcium binding protein a10
SBP: systolic blood pressure
*SCG2*: secretogranin ii
*SCNN1B*: sodium channel epithelial 1 subunit beta
*SCNN1G*: sodium channel epithelial 1 subunit gamma
*SERTAD2*: serta domain containing 2
SNS: sympathetic nervous system
SP: São Pedro (quilombo population)
*TENM4*: teneurin transmembrane protein 4
TF: transcription factor
TU: Sapatu (quilombo population)
YRI: Yoruba in Ibadan, Nigeria
*ZZEF1*: zinc finger zz-type and ef-hand domain containing 1

