## Supplementary material for "Genomic Exploration of Essential Hypertension in African-Brazilian Quilombo Populations: A Comprehensive Approach with Pedigree Analysis and Family-Based Association Studies": Supp. Material

### METHODS

#### DATA S1: SAMPLES AND SNP GENOTYPING

Individuals underwent physical examinations, including weight, height, and blood pressure measurements, all conducted by trained physicians. Weight and height measurements were taken with participants in lightweight clothing and without shoes. Blood pressure was assessed using either a portable monitor (Omron) or a sphygmomanometer. We took two systolic and diastolic pressure measurements, calculating the average of each. For participants examined multiple times, we considered the most recent measurements. These measurements were taken three times for each participant, and the mean of the three measurements was calculated using standardized methods[136].

Blood samples were processed for DNA extraction using one of the three strategies: (1) the standard phenol and chloroform techniques, (2) the Gentra Systems Autopure LS equipment (Qiagen, Limburg-Netherlands), or (3) the DNA extraction kit QIAsymphony DSP DNA Midi Kit (Qiagen, Limburg-Netherlands). DNA samples were quantified (NanoDrop ND-1000 spectrophotometer / Thermo Fisher Scientific) and prepared according to Affymetrix (Santa Clara, CA, USA) instructions (15 ng/µL, 50µL, OD260/OD280 ratio 1.8-2.0 and OD260/OD230 ratio >1.5).

DNA samples (n=431) were subjected to SNP genotyping by the Affymetrix Microarray Research Services Laboratory (Santa Clara, CA, USA) using the Axiom® Genome-Wide Human Origins 1 Array SNPs. Raw data were processed using Axiom Analysis Suite 5.1.1 software, annotated for genome build GRCh37/hg19, and subjected to quality control using 22 different parameters from the Affymetrix human v.5a threshold[58]. The genotype data were then converted to PLINK format[64].

From the interview data collected from each participant, we constructed six pedigrees using GenoPro 2018 v.3.0.1.4 software. These pedigrees encompassed individuals from eight different populations (Figure 2) and included 431 typed samples and 673 untyped samples, which were used to establish family links. Each sample presented individual-specific code, parental codes, gender information, phenotype information and genotypes for each marker (ACTG as allele coding). Each dataset coherently represented family relationships observed in the pedigree. For it, we handled the files as PLINK binary biallelic genotype format (.bim, .bed, and .fam). We validated the pedigree structures through pairwise kinship coefficients ($\Phi$) with the assistance of KING-robust v.2.2.5[60], Checkped (MORGAN v.3.4 suite v.34[61,62]), and Relationship Check (PBAP v.1[63]) software.

To detect genotyping errors, we conducted further quality control with the assistance of KING v.2.2.5 [60] and PLINK v.1.90b[64] software. We excluded samples with a genotyping rate of <95%, SNPs with a genotyping rate of <95%, monomorphic SNPs, and SNPs causing heterozygous haploid calls across all remaining individuals. SNPs not in Hardy-Weinberg equilibrium ($p < 1\times10^{-3}$) were also removed. We updated SNP identification from the Affymetrix default pattern (AX-ID) to the dbSNP standard (rsID) using Affymetrix annotation files (GRCh37/hg19) and provided a reliable genetic location (cM) to datasets using Rutgers Combined Linkage-Physical Map v.3[65].

DATA S2: ANCESTRY ESTIMATION

The next stage was to obtain estimates of local ancestry fractions. The execution of this approach also allowed us to determine individual-specific and pedigree-specific ancestry. To perform this task, we established a reference dataset using 1000 Genomes Project phase 3[137] and Stanford HGDP SNP Genotyping[138] data. This dataset was composed of 189 samples, as follows: 63 European samples (CEU population; Northern Europeans from Utah), 63 African samples (YRI population; Yoruba in Ibadan, Nigeria) and 63 Native American samples (Colombian, Maya, and Pima populations). The inference dataset comprised samples from 431 individuals from the populations of quilombos remnants.

We gathered common SNPs between reference and inference datasets. Additionally, SNPs not on the reference strand were reported by SNPFlip script[139] using data from 1000 Genomes Project as a reference and flipped using PLINK 1.90b[64]. Then, we merged the reference and inference datasets into a single dataset and filtered for missingness per marker (≤ 95% genotyping rate) by PLINK 1.90b[64]. We aligned the dataset using 1000 Genomes Project phase 3 data and inferred the haplotypes using SHAPEIT2 software[68], applying duoHMM method.

Lastly, we converted the phasing output files to RFMix[69] input files according to specific Python scripts[140]. Local ancestry calls were estimated by RFMix software. Global ancestries fractions for each sample and pedigree ancestries were estimated from RFMix output files.

DATA S3: PEDIGREE (LINKAGE) ANALYSIS

MORGAN power resides in the Markov Chain Monte Carlo (MCMC) approach that is explored, a sampling method more versatile than exact computation based-approaches, such as Elston-Stewart and Lander-Green algorithm. Since it is based on MCMC, MORGAN can handle many markers simultaneously with a large number of individuals on each pedigree.

DATA S3.1: Marker Subpanels Selection

The first step to perform linkage analysis using MORGAN suite is to acquire pedigree datasets files into transpose format (.tpedo, .mped, .tphen, .tgen, .tmap, .tind, .tfam and .tped). We handle this aided by transpose_fileset script (PBAP2 software). To achieve this, we built pedigree, phenotype, marker map, and genotype files using in-house developed R scripts.

We designed linkage analysis to be performed triple and independently. The goal here is to provide independent results that can be compared and corroborated. From the complete set of markers (for each pedigree), we carefully selected and divided them into 3 non-overlapping subsets (or sub dataset, or subpanels) of markers. This task was performed by marker_subpanels script (PBAP2 software) using “transpose_fileset” output files. We selected the markers according to the following criteria/parameters:

a) maximum LD threshold (r2 = 0.04);
b) MAF minimum and maximum cut-offs, and gap filling decrement value (0.2 0.5 0.05);
c) monomorphic markers (all excluded);
d) minimum intermarker distance (MID; cM), window size (cM) for LD-based SNP pruning and gap filling iteration increment factor (0.5 1.0 0.1);
e) source of reference populations (1000 Genomes Project phase 3);
f) number of marker subpanels (3);
g) marker completion threshold (90%).

Inherent in the design of this approach is a quality difference between the markers assigned to each subpanel. The first marker subpanel is the best one (gold-standard), which contains the markers that best match each selection parameter and is used for the discovery analyses. The additional subpanels also included good markers, however, since there was no overlap among different subpanels and we had already selected the best markers for the first one, we may expect a slight decrease in quality for subsequent subpanels, which is not sufficient to compromise the linkage analysis results. These additional marker subpanels are also used for linkage analysis.

DATA S3.2: Inheritance Vectors (or Meiosis Indicators)

The descent through the pedigree of genes at marker and trait locus is tracked by inheritance vectors (IVs), also known as meiosis indicators[141]. IVs are a set of FGLs — or founder genome labels (unique identifiers assigned to each of the two haploid genomes of each founder) — that specify the flow of founder alleles in a pedigree. IVs represent the flow of chromosomes through pedigrees. At each locus, non-founders are assigned two representing genes inherited from the individual's father and mother. The IBD (identity by descent) graph synthesizes the inheritance pattern. The inheritance vectors represent the IBD pattern of a gene among observed individuals, which is critical to computing observed trait probability over linkage analysis.

DATA S3.3: LOD Score Calculation

The LOD scores were estimated using MORGAN v3.4 suite (Figure 1H and 1J) following a two-step approach: IVs sampling using gl_auto software and LOD score calculations using gl_lods software. Both steps were pedigree-specific.

For the IVs estimation, we used:

a) pedigree data;
b) genotype data;
c) allelic frequencies;
d) list of markers of interest
e) number of MCMC iterations (200,000-600,000);
f) percentage of burn-in iterations (20% of MCMC iterations);
g) thinning factor (25-100);
h) IV realizations (1,000-6,000);
i) maximum number of IBD graphs per component (1,000-3,000) to compute IVs pedigree- and subpanel-specific (as .fgl format file).

At first, IVs were estimated only for the first subpanel (the best one, which contains the markers that most meet each one of the selection parameters) using alternate SNP markers (2,500-3,000 SNPs). This strategy is called "linkage analysis genome-wide scan" (also known as "scan"). The computation complexity to estimate the IVs is directly related to the pedigree structure, and the computation time is linear with the number of markers. So, to optimize the computational time, we opted to run the "scan" analysis using a thinned sparse subpanel (50% of all the subpanel 1 available SNP markers); we used the original dense marker subpanel in a second round of analyses (detailed below). This strategy proved to be efficient in reducing the time to sample the IVs; the computation time to run gl_auto by pedigree using the thinned subpanel ranged from 21 to 25 days while using the original subpanel, the analyses took from 22 to 49 days, varying with the size of the chromosome.

We used the IVs sampled in the previous section and the pedigree data to compute the LOD scores. The gl_lods parameters used were:

a) IBD graphs number (1,000~3,000);
b) list of markers of interest;
c) trait allele frequencies (0.95~0.99 and 0.01~0.05, for alleles 1 and 2, respectively);
d) data class as discrete;
e) trait incomplete penetrance rate as 70% (0.05 0.7 0.7 for trait locus genotypes [1,1], [1,2], [2,2] respectively);
f) base log likelihood unique by pedigree (used to normalize the IBD-based LOD score computed by gl_lods).

The regions presenting peak LOD score ≥ 1 (loosely defined) for the "scan" analysis results were considered candidates. Each chromosome containing these candidate regions was integrally submitted to linkage "dense" analyses, as follows. The linkage "dense" analysis was performed independently for the 3 subpanels. For this, IVs were again estimated via gl_auto software (parameters presented above), this time using all the available SNP markers for the entire chromosome containing the candidate region. Subsequently, LOD scores were again computed via gl_lods software (parameters presented above). Finally, we considered as regions of interest (ROIs) the ones reaching maximum LOD score ≥ 1.50 (loosely defined, 5% of tolerance) for at least one of the 3 subpanels, with mandatory compliance for subpanel 1 (obligatory positive result without deviating over 37%). The ROI boundaries were defined based on the position of the markers presenting LOD score ≥ 1 (loosely defined). Lastly, we investigated all the ROIs by fine-mapping strategies.

DATA S3.4: MCMC (Markov chain Monte Carlo) Convergence Diagnostic Analysis

MORGAN is a Markov chain Monte Carlo-based programs suite which uses sampling to average a mapping statistic over many such samples. As with any other stochastic method, one must ensure that proper convergence is achieved in the sampling process. Therefore, we implemented a diagnostic analysis of the MCMC runs helping to determine the adequate run conditions.

Ideally, the MCMC algorithms would be run for a large number of iterations (cycles of MCMC sampling) to ensure the convergence (when the sampling algorithm converges to a stationary distribution equal to the target distribution) and all realizations of S (set of IVs) would be used in the LOD score computation. In practice, only a subset of realizations is used. Hence, the MCMC run conditions should be assessed to guarantee the chain convergence and accurate results, such as: the number MCMC iterations (total number of main sampling iterations), burn-in period (initial realizations to be discarded to avoid the possible bias caused by the effect of starting values), and saved realizations (frequency with which to compute the contributions to the location LOD scores). We evaluated the MCMC run conditions (Figure S1) under three different graphic tools.

MCMC run length: We got the number of saved realizations (SR) and computed SR/100 LOD score vectors using 100 sequentially large subsets of realizations. If the run is long enough, additional realizations substantially affect the LOD score, creating an unnecessary computational burden. If it is short, it will not converge (Figure S1A); Autocorrelation of LOD scores: when the MCMC algorithm performs well, we do not see highly autocorrelated MCMC samples. The magnitude of the autocorrelations is small, providing evidence of good mixing of the chain (Figure S1B); MCMC run stability: when the MCMC algorithm performs well, the LOD score signals are relatively stable across different portions of the space of realizations (Figure S1C-D)[142].

Getting an accurate result requires testing and personalization. Therefore, we tuned the run conditions in the smaller chromosome with evidence of positive LOD score for each pedigree. The effect of the pedigree structure was the same across chromosomes, so that the inference about the run conditions done for one chromosome can be safely applied for all chromosomes in the same pedigree.

Each tunning round — for better MCMC diagnostics — consisted of new IV calculations, new LOD score computations and graph plotting for MCMC diagnostics (as presented below). The IVs and LOD scores were estimated as previously described. The graph plotting required inheritance vectors, pedigree data, IV realizations number, markers number, IBD graphs number and a tweaked setup. These tweaks were different for each round and pedigree-specific. We started from an initial setup (100,000 MC iterations, 40,000 burn-in iterations, 25 saved realizations), and tested dozens of possible combinations until we got tunned setup for each pedigree based on the convergence assessment.

Figure S1. MCMC run diagnostics

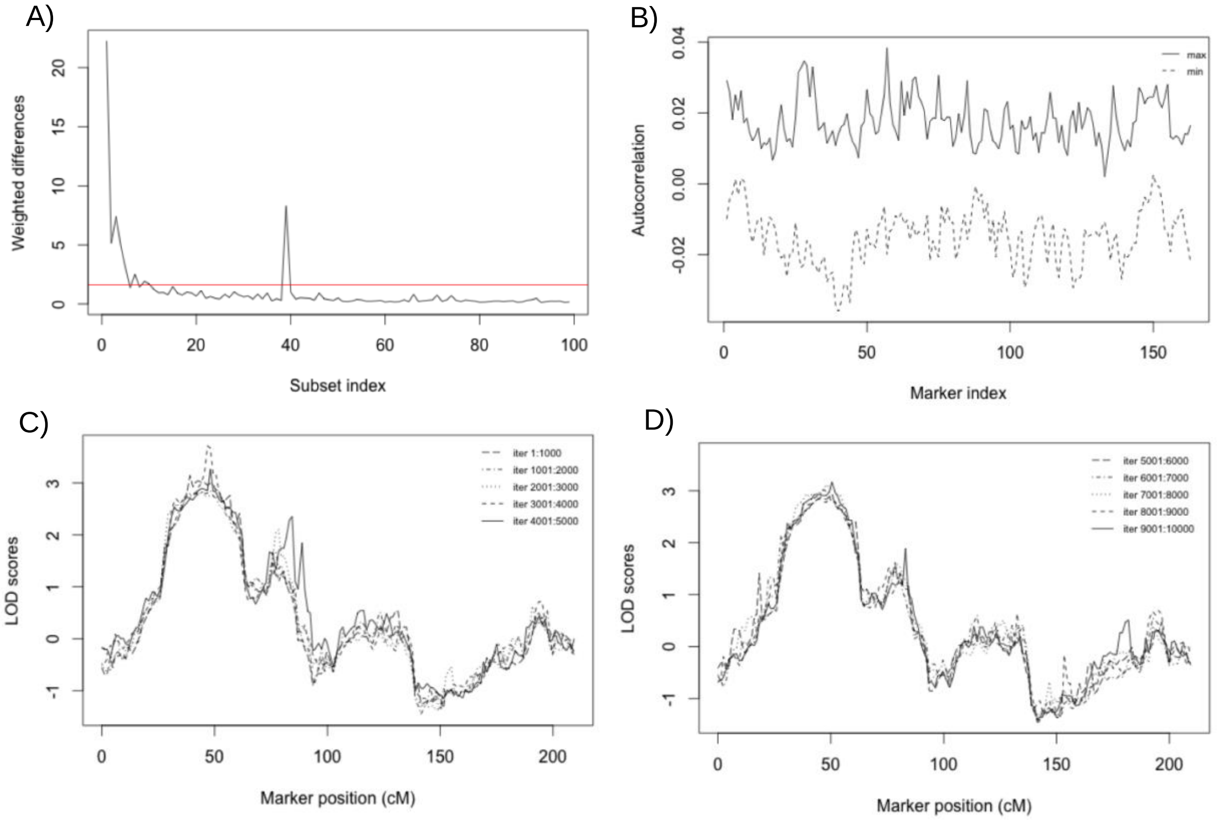


Once we had found the proper setup for each pedigree, we re-executed genome-wide scan and dense analysis, redoing inheritance vectors estimation and LOD score calculation.

Here will be presented and discussed the diagnostic for MCMC runs and the final setup for the linkage analysis. The MCMC runs diagnosis aimed the smallest chromosome with a positive LOD score (to optimize computational processing time) by each pedigree. This process consisted of a series of MCMC testing-adjusting rounds, and for every round the numbers of MC iterations, burn-in iterations and saved realizations were adjusted. Each round included: 1º) parameter adjustment; 2º) inheritance vector calculations; 3º) LOD score calculations; 4º) plotting graphs; and 5º) results analysis. Once the ideal configuration was reached, the parameters were applied to the complete set of chromosomes.

The MCMC setup started from a default set arbitrarily defined for all pedigrees: 100,000 MC iterations, 40,000 burn-in iterations, 25 saved realizations, 1,000 ibdgraphs and output scores saved at every 25 scored MC iterations. Parametric linkage analysis was under an autosomal-dominant model with the following parameters: a risk allele frequency of 0.01, an incomplete penetrance of 0.70 (for genotypes with 1 or 2 copies of the risk allele), and a phenocopy rate of 0.05. This setup was specified considering the average complexity of all pedigrees (samples and meiosis number) and the disease heritability pattern.

For each pedigree we achieved an optimized MCMC setup used for IV estimations.

**Abobral (ABDR) Pedigree**

For ABDR pedigree, 5 round tests were performed until the following result (Figure S2) was obtained. The optimized MCMC setup is presented below:

a) 150,000 MC iterations;
b) 30,000 burn-in iterations;
c) 3,000 ibdgraphs;
d) Output scores every 25 scored MC iterations;
e) 0.05 risk allele frequency;
f) 0.05 0.70 0.70 trait incomplete penetrance (genotypes [1,1], [1,2], [2,2]).

Figure S2 – MCMC run diagnostics for ABDR pedigree

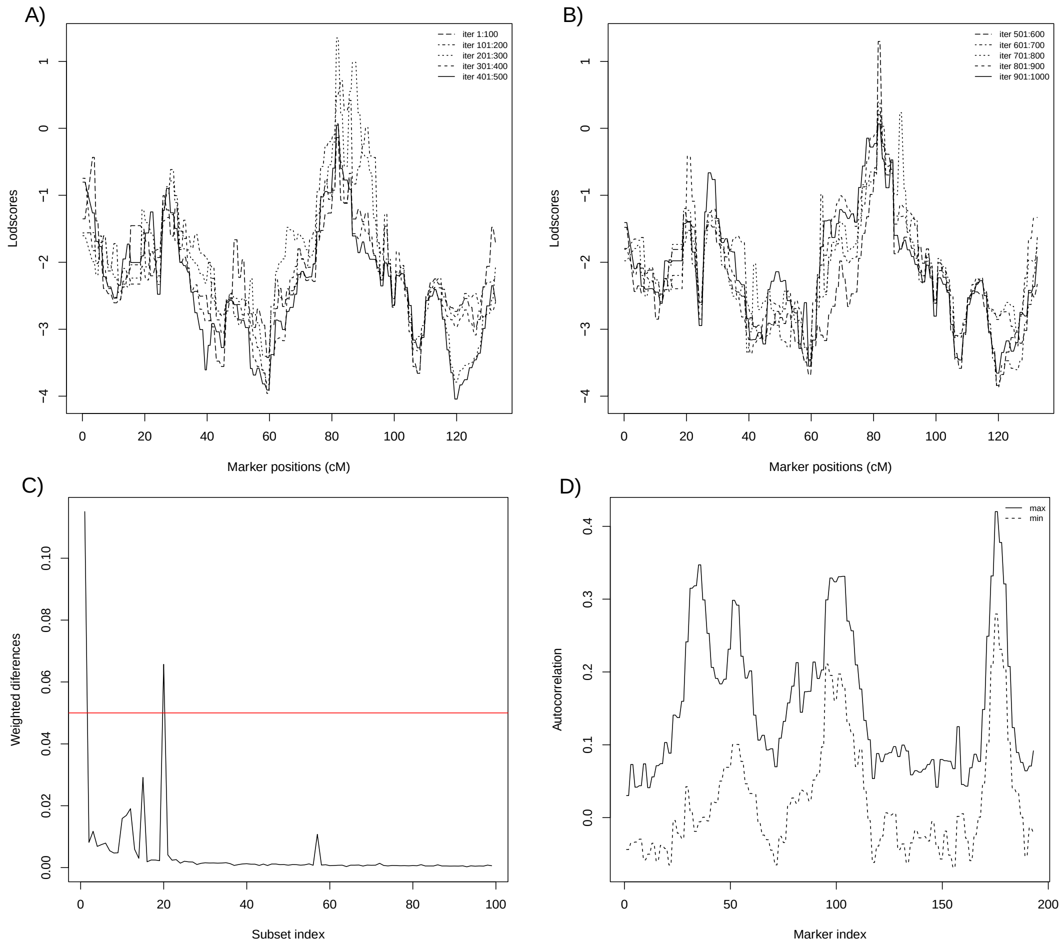


A) LOD score vectors computed for each 100 saved realizations of S.; B) Continuation of A figure, until 1,000 saved realizations; C) Weighted differences between LOD score vectors computed using adjacent sequential subsets of S: increment of 100, 100 subsets in total. Red line represents a threshold for convergence; D) The maximum and minimum of the first 10 lag autocorrelations.

**André Lopes and Nhunguara (ANNH) Pedigree**

For ANNH pedigree, 20 round tests were performed until the following result (Figure S3) was obtained. The optimized MCMC setup is presented below:

a) 250,000 MC iterations;
b) 50,000 burn-in iterations;
c) 3,000 ibdgraphs;
d) Output scores every 25 scored MC iterations;
e) 0.05 risk allele frequency;
f) 0.05 0.0.5 0.70 trait incomplete penetrance (genotypes [1,1], [1,2], [2,2]).

Figure S3 – MCMC run diagnostics for ANNH pedigree

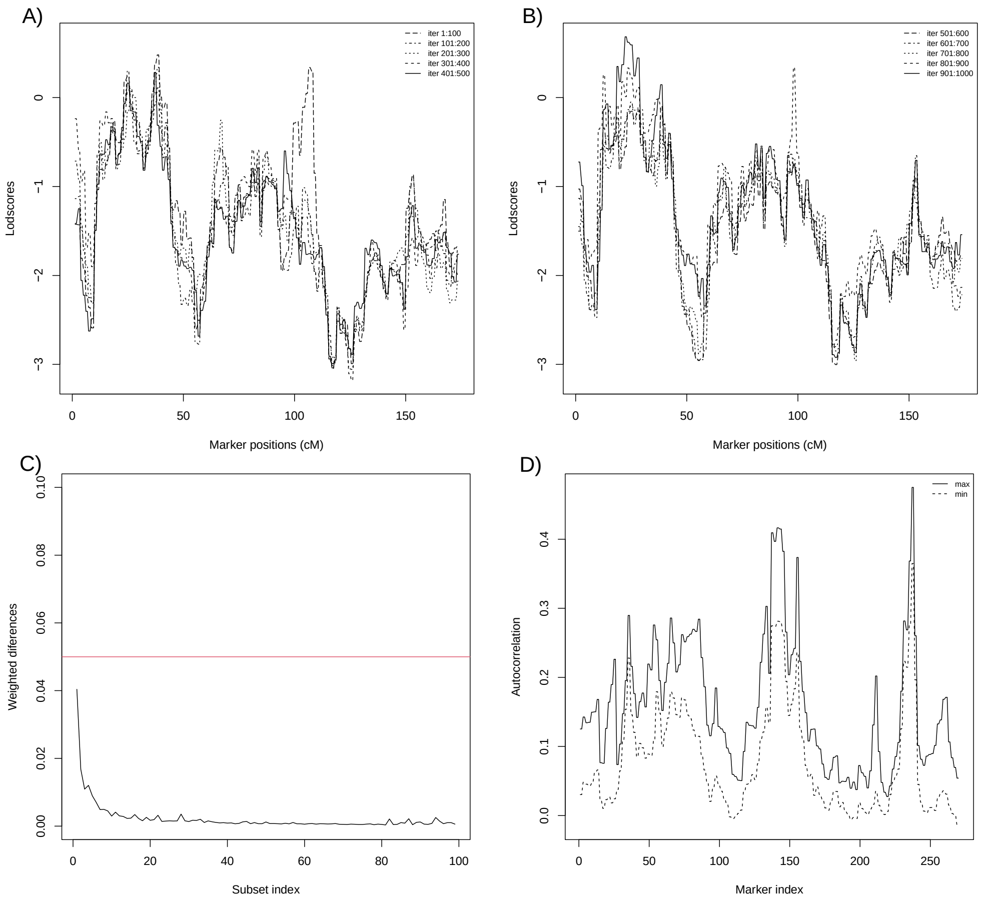


A) LOD score vectors computed for each 100 saved realizations of S.; B) Continuation of A figure, until 1,000 saved realizations; C) Weighted differences between LOD score vectors computed using adjacent sequential subsets of S: increment of 100, 100 subsets in total. Red line represents a threshold for convergence; D) The maximum and minimum of the first 10 lag autocorrelations.

#### Galvão and São Pedro (GASP) Pedigree

For GASP pedigree, 20 round tests were performed until the following result (Figure S4) was obtained. The optimized MCMC setup is presented below:

a) 500,000 MC iterations;
b) 100,000 burn-in iterations;
c) 6,000 ibdgraphs;
d) Output scores every 100 scored MC iterations;
e) 0.05 risk allele frequency;
f) 0.05 0.0.70 0.70 trait incomplete penetrance (genotypes [1,1], [1,2], [2,2]).

Figure S4 – MCMC run diagnostics for GASP pedigree

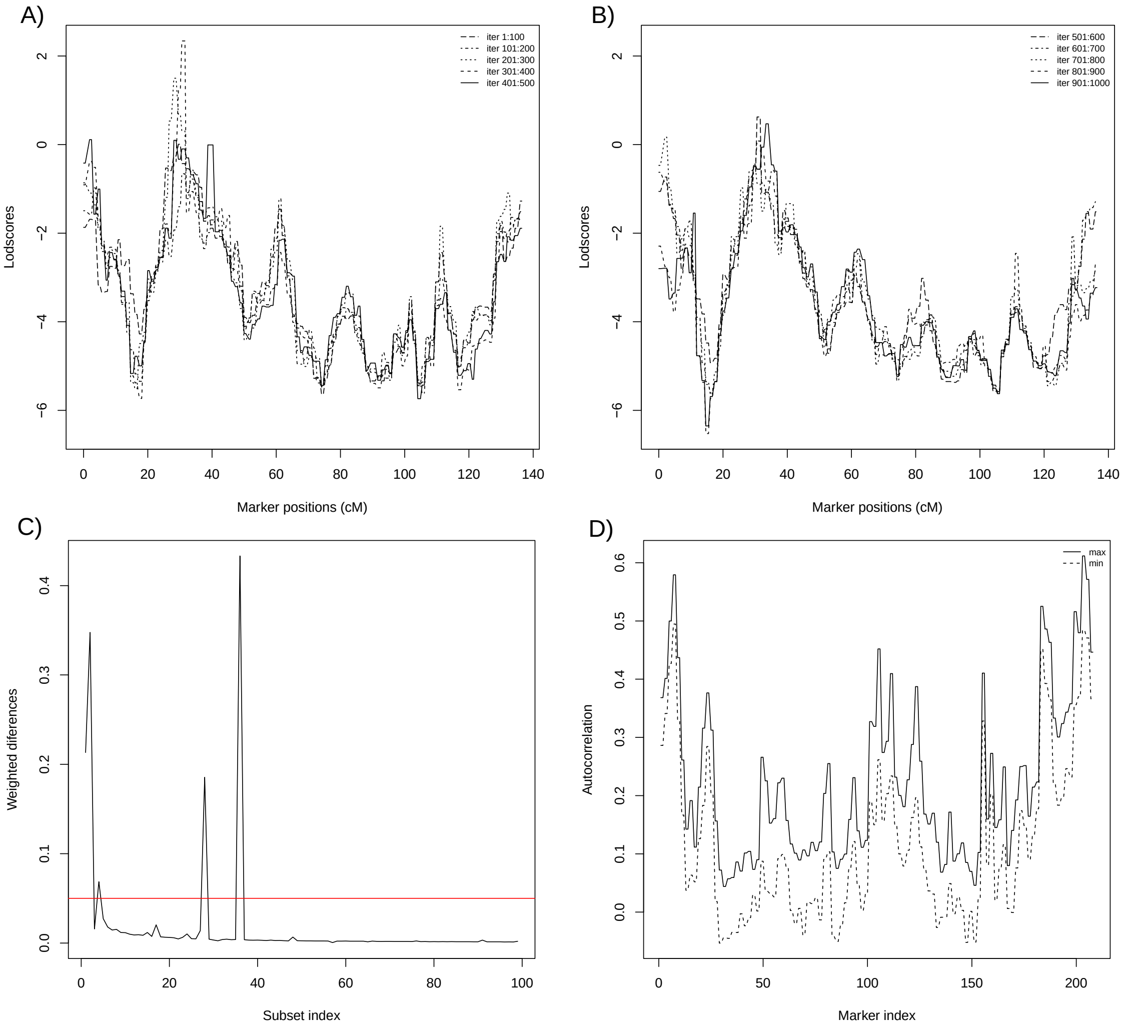


A) LOD score vectors computed for each 100 saved realizations of S.; B) Continuation of A figure, until 1,000 saved realizations; C) Weighted differences between LOD score vectors computed using adjacent sequential subsets of S: increment of 100, 100 subsets in total. Red line represents a threshold for convergence; D) The maximum and minimum of the first 10 lag autocorrelations.

**Ivaporunduva (IV) Pedigree**

For IV pedigree, 7 round tests were performed until the following result (Figure S5) was obtained. The optimized MCMC setup is presented below:

a) 150,000 MC iterations;
b) 30,000 burn-in iterations;
c) 3,000 ibdgraphs;
d) Output scores every 25 scored MC iterations;
e) 0.01 risk allele frequency;
f) 0.05 0.0.70 0.70 trait incomplete penetrance (genotypes [1,1], [1,2], [2,2]).

Figure S5 – MCMC run diagnostics for IV pedigree

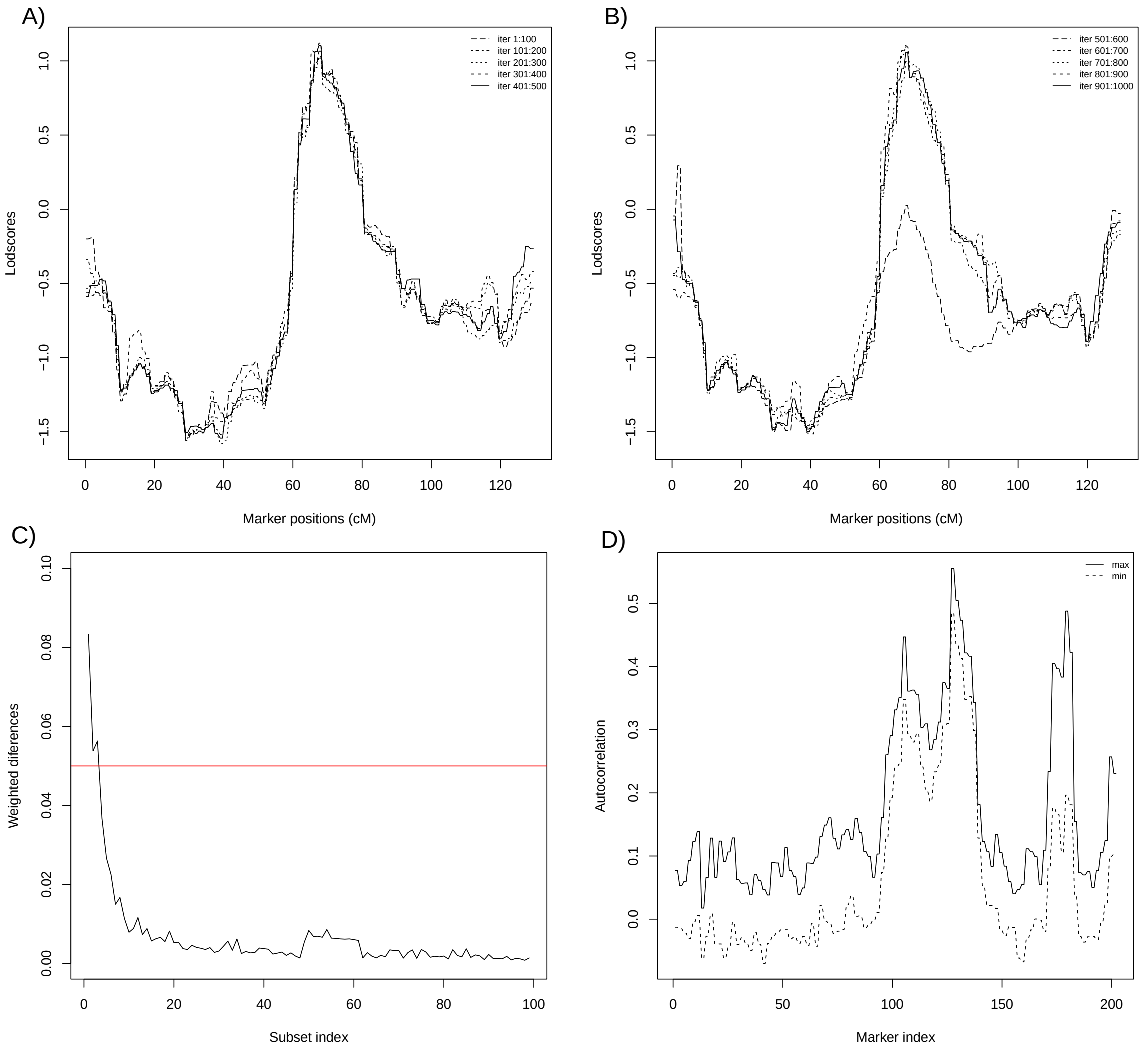


A) LOD score vectors computed for each 100 saved realizations of S.; B) Continuation of A figure, until 1,000 saved realizations; C) Weighted differences between LOD score vectors computed using adjacent sequential subsets of S: increment of 100, 100 subsets in total. Red line represents a threshold for convergence; D) The maximum and minimum of the first 10 lag autocorrelations.

**Pedro Cubas (PC) Pedigree**

For PC pedigree, 13 round tests were performed until the following result (Figure S6) was obtained. The optimized MCMC setup is presented below:

a) 200,000 MC iterations;
b) 40,000 burn-in iterations;
c) 3,000 ibdgraphs;
d) Output scores every 100 scored MC iterations;
e) 0.01 risk allele frequency;
f) 0.05 0.0.70 0.70 trait incomplete penetrance (genotypes [1,1], [1,2], [2,2]).

Figure S6 – MCMC run diagnostics for PC pedigree

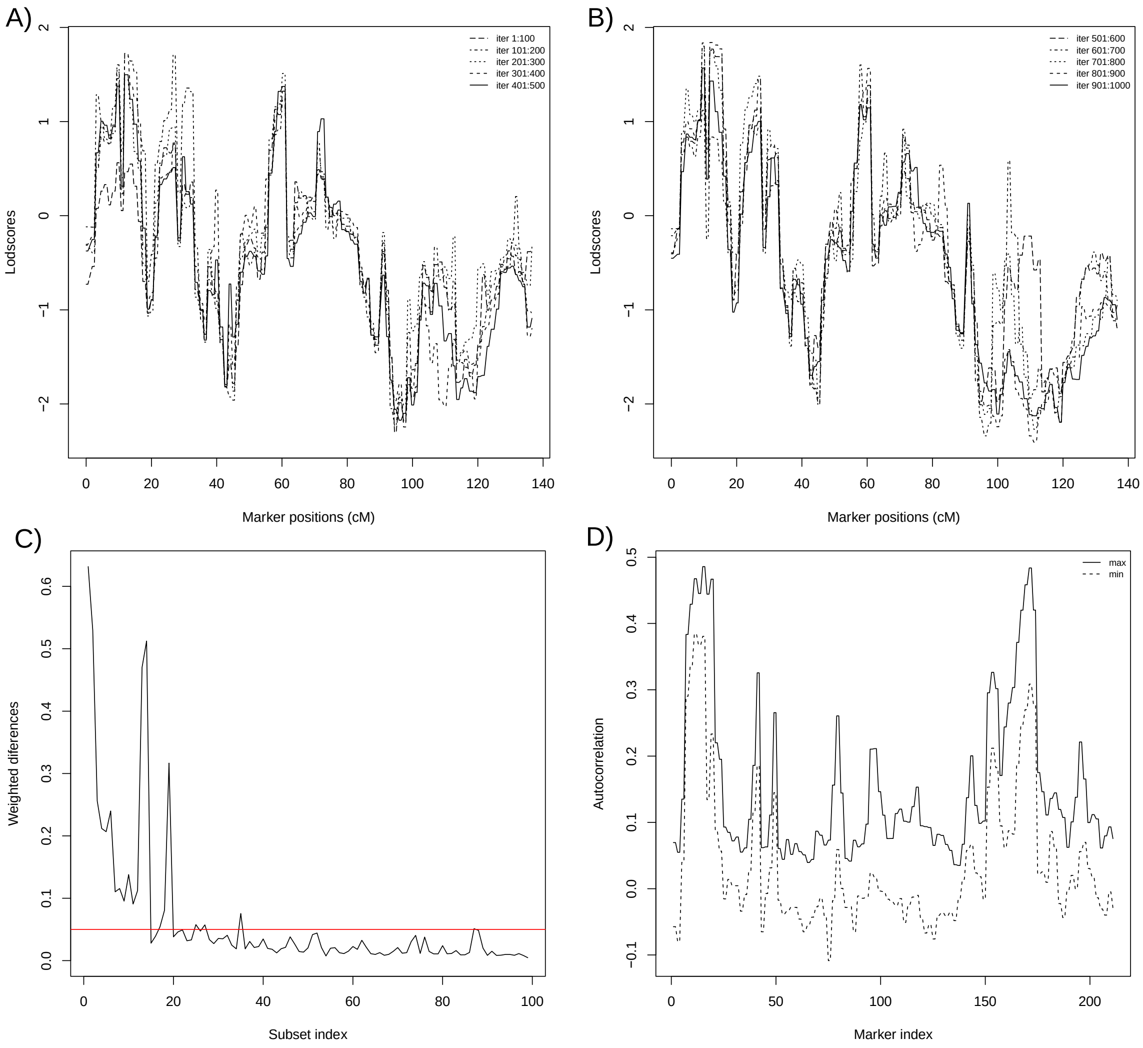


A) LOD score vectors computed for each 100 saved realizations of S.; B) Continuation of A figure, until 1,000 saved realizations; C) Weighted differences between LOD score vectors computed using adjacent sequential subsets of S: increment of 100, 100 subsets in total. Red line represents a threshold for convergence; D) The maximum and minimum of the first 10 lag autocorrelations.

**Sapatu (TU) Pedigree**

For TU pedigree, 9 round tests were performed until the following result (Figure S7) was obtained. The optimized MCMC setup is presented below:

a) 200,000 MC iterations;
b) 40,000 burn-in iterations;
c) 3,000 ibdgraphs;
d) Output scores every 100 scored MC iterations;
e) 0.01 risk allele frequency;
f) 0.05 0.0.70 0.70 trait incomplete penetrance (genotypes [1,1], [1,2], [2,2]).

Figure S7 – MCMC run diagnostics for TU pedigree

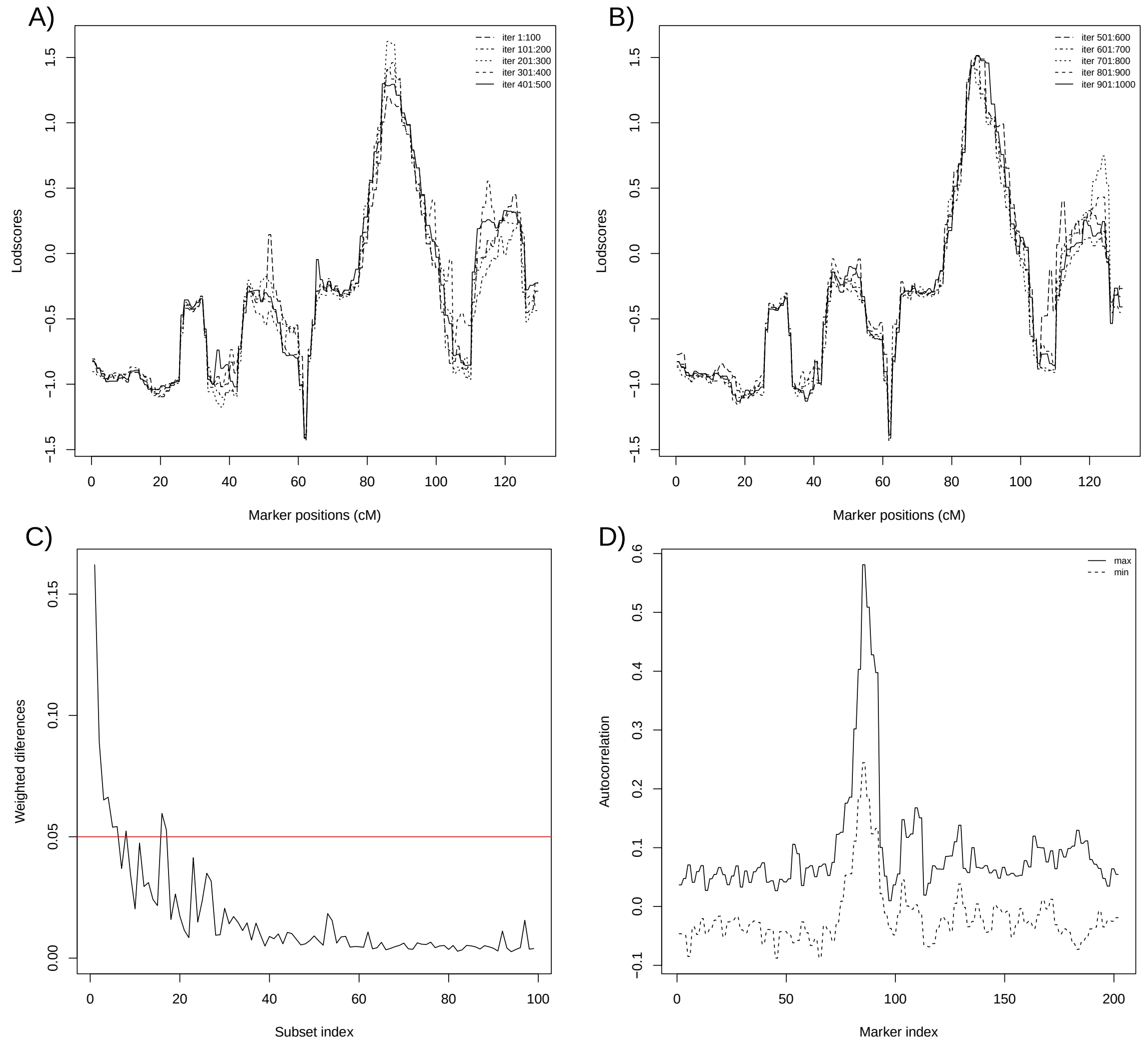


A) LOD score vectors computed for each 100 saved realizations of S.; B) Continuation of A figure, until 1,000 saved realizations; C) Weighted differences between LOD score vectors computed using adjacent sequential subsets of S: increment of 100, 100 subsets in total. Red line represents a threshold for convergence; D) The maximum and minimum of the first 10 lag autocorrelations.

DATA S4: Family-Based Association Studies

DATA S4.1: PCA and GRM Estimation

First, we implemented principal components analysis (PCA) and prepared downstream necessary files as the genetic relationship matrix (GRM). Since the PCA requires knowing if there is a relationship between samples, the initial estimates of pairwise kinship coefficients (Φ) were calculated by KING-robust v.2.2.5[60], and the output (.kin and .kin0) was converted to matrix format by using GENESIS v.2.2.2 R package[57]. KING-robust is robust to population structure but not to admixture. The matrix is used for partitioning the sample into the ‘unrelated’ and ‘related’ subsets. Before running KING-robust and ensuring the result, all sample relatedness was double-checked by Checkped (MORGAN v.3.4 suite) and Relationship Check (PBAP v.1[63]) software. The next step was to run the PC-AiR function (GENESIS v.2.2.2 R package), which serves to detect population structure in a sample (accounting for sample relatedness to provide accurate ancestry inference not confounded by the family structure). It is done by selecting a set of unrelated samples that is maximally informative about all ancestries in the sample. We used this unrelated set for PCA, then projected the relatives onto the PCs. In the first iteration (1º round), we used the KING estimates for both kinship and ancestry divergence. KING kinship estimates were negative for samples with different ancestry. It was also required: genotype data (from inference and reference dataset as .gds file converted by the SNPRelate package v.1.6.4 from PLINK format including the 620 samples) and parameters for the following setup: 140,283 SNPs used; 0.025 as kinship threshold and -0.025 as divergence threshold; MAF = 0.05; PCs to return = 10.

The first 4 PCs (PC-AiR output) separated populations, so we used them to compute kinship estimates adjusting for ancestry using the PC-Relate function (GENESIS v.2.2.2 R package[57]). To run PC-Relate, we used: the PC-AiR output; genotype data, with SNPs organized by iterators blocks by GWASTools v.1.18.0 R package[143]; MAF = 0.05; and unrelated samples as the training set. The PC-Relate output (GRM) are ancestry-adjusted kinship estimates, and once we have them, we use them to better adjust for ancestry in the PCs, rerunning the PC-AiR function.

In the second iteration (2º round), we ran the PC-AiR function using PC-Relate GRM for kinship and KING estimates for ancestry divergence, and the same parameters used previously. Then we used the revised PCs (the PC-AiR’s 2º round output) to compute new kinship estimates rerunning the PC-Relate function to obtain a new GRM (the PC-Relate’s 2º round output). After that we calculated the variation by top 10 PCs (chosen based on Kaiser’s rule) and plotted the first four.

DATA S4.2: Imputation Strategies

To perform pedigree-based imputation, we used GIGI2 software. GIGI2 is a computationally efficient approach for imputing dense genotypes in large pedigrees, which accompanies the original GIGI software. This approach to imputation uses a sparse set of “framework markers” genotyped on most subjects plus a set of “dense markers” genotyped on a few subjects. The imputation relies on a correlation resulting from inheritance in pedigrees through the inheritance of shared chromosome segments as represented by IVs[144]. For these computations, we used each pedigree, dense markers (all available markers), IVs and framework markers (subpanels markers) data. We set up the following parameters: iterations = 2,000; genocall = 1. We checked imputation accuracy by evaluating Pearson’s squared correlation ($R^2$) over R script cor function.

To perform population-based imputation, we first pruned markers based on missing rate using Plink1.90b[64]. Later we separated the dataset by chromosomes and estimated the haplotypes (phasing). The phasing process has counted on the flipping of SNPs not on the reference strand by SNPFlip. Then we checked the alignment between inference data and reference data (1000 Genomes Project phase 3) and phased by using the software packages SHAPEIT2+duoHMM[68]. Later we ran MINIMAC4 software to perform population-based imputation. We used SHAPEIT2+duoHMM output files as inference dataset, 1000 Genomes Project phase 3 as reference dataset and set up the following parameters: window = 500,000,000; rounds = 20; states = 400; allTypedSites; CPUs = 18; vcfBuffer = 480. We checked imputation accuracy by evaluating Pearson's squared correlation ($R^2$) over bcftools v.1.12 software.

#### DATA S4.3: Performing Association Testing

We performed association testing independently for each variants datasets resulting from the two imputation strategies, pedigree-based and population-based, for each ROI. Unlike linkage analyses — performed on a pedigree-restricted basis — the association studies were performed cross-sectional to all samples, thus investigating each ROI overall individual data (431 samples) from all pedigrees (6 pedigrees) to gain analysis power.

We developed all further steps coding in R script. For each of the 22 ROI we loaded the dataset (containing the 431 samples) in .gds format. We handled this file by using GWASTools v1.18.0 R package[143] to get block iterator (that reads variants in blocks). We used Genetic Type I Error Calculator software v.1.0[145] to calculate the effective number of independent markers ($M_e$) and to adjust both suggestive and significant p-value thresholds for multiple testing. The single-variant association tests utilized the assocTestSingle function (GENESIS v.2.20.1 R package[57]), implementing the adjusted GLMM to perform Score tests, obtaining the value of the score function, the estimated standard error of the score, the score Z test statistic, p-value, the effect size estimate for each additional copy of the effect allele and the proportion of phenotype variance explained. We also evaluated the genomic inflation factor, which estimates the amount of inflation by comparing observed test statistics across all genetic variants to those expected under the hypothesis of no effect[146] — by estimating lambda ($\lambda$).

#### DATA S5: Investigation of EH-related Genes

The goal of this step was to find out which genes identified in the ROI are related to hypertension. We first organized each ROI in a database and used biomaRt R package[147] to investigate all possible genes present in these ROIs. Then we gathered all "essential hypertension" or "high blood pressure" related genes on several databases: NCBI PubMed[81], MedGen[90], MalaCards[91], ClinVar[83], Ensembl BioMart[92], and GWAS Catalog[93]. These lists of essential hypertension-related genes were paired and crossed with all-found genes by candidate region. We listed the matched ones in the database, annotating them with genomic location (GRCh37/hg19), expression, and publication data: physical location (bp), cytogenetic band, summary, molecular function, related phenotype, gene ontology and description. Additionally, we ranked all matched genes through VarElect[94] based on the VarElect score (connection strength between genes and phenotype), average disease-causing likelihood and global ranking (position of each of those matched genes within a global ranking of phenotype-related genes) to prioritize.

RESULTS

DATA S6: Pedigrees

Pedigrees were built from a dataset of 1,104 samples, 431 genotyped using the SNPs Axiom® Genome-Wide Human Origins 1 Array, and another 673 non-genotyped included to compose the pedigree structure. The individuals were organized into 6 pedigrees corresponding to the quilombos remnant populations of Abobral (**ABDR**), André Lopes and Nhunguara (**ANNH**), Galvão and São Pedro (**GASP**), Ivaporunduva (**IV**), Pedro Cubas (**PC**), Sapatu (**TU**). The pedigree drawing was achieved based on family history and reflects the kinship coefficients Φ, performed according to the methodology proposed.

**Table S1. Age differences across pedigrees in affected and unaffected individuals.** Comparison of mean ages between unaffected and affected individuals across different pedigrees. The table displays mean age ± standard deviation for each group, along with t-values, degrees of freedom (df), p-values, and 95% confidence intervals for the difference in mean ages. Statistically significant differences are observed across all pedigrees, with affected individuals consistently showing higher mean ages compared to unaffected individuals.

| Pedigree | Mean Age (Unaffected) | Mean Age (Affected) | t-value | df | p-value | 95% Confidence Interval |
|---|---|---|---|---|---|---|
| **ABDR** | 35.9 ± 15.9 | 49.7 ± 19.5 | 3.20 | 64.6 | $2.112343 \times 10^{-3}$ | [5.22, 22.5] |
| **ANNH** | 36.4 ± 16.7 | 57 ± 16.6 | 5.81 | 77.9 | $1.291263 \times 10^{-7}$ | [13.5, 27.7] |
| **GASP** | 37 ± 13.5 | 54.3 ± 16.8 | 5.46 | 84.2 | $4.771149 \times 10^{-7}$ | [11.0, 23.6] |
| **IV** | 33.2 ± 14.1 | 64.3 ± 12.7 | 7.08 | 21.4 | $4.963544 \times 10^{-7}$ | [22.0, 40.2] |
| **PC** | 38.4 ± 14.6 | 60.6 ± 17 | 4.71 | 22.4 | $1.026268 \times 10^{-4}$ | [12.4, 31.9] |
| **TU** | 38.2 ± 17.2 | 53.8 ± 15.1 | 3.60 | 38.7 | $8.866232 \times 10^{-4}$ | [6.84, 24.3] |
| **TOTAL** | 55.11 ± 17.3 | 36.73 ± 15.4 | 11.2 | 323.7 | $6.855379 \times 10^{-25}$ | [15.15, 21.60] |

**Table S2. Ancestry proportions among Brazilian regions.** Ancestry estimates across Brazilian regions, ranked by North, Northeast, Center-West, Southeast, and South. The data includes the number of samples analyzed, ancestry proportions, and corresponding study source.

| Brazilian Region | Sample size (N) | Ancestry Proportions | | | Study Source |
|---|---|---|---|---|---|
| | | African | European | Native American | |
| **North** | 3,092 | 19.8% | 52.6% | 27.7% | de Souza et al., 2019[50] |
| | 368 | 17% | 51% | 32% | Saloum et al., 2013[148] |
| | 203 | 10.5% | 68.8% | 18.5% | Pena et al., 2011[149] |
| **Northeast** | 1,309 | 50.8% | 42.9% | 6.4% | Kehdy et al., 2015[150] |
| | 7,837 | 35.2% | 50.8% | 13.9% | de Souza et al., 2019[50] |
| | 237 | 28% | 56% | 16% | Saloum et al., 2013[148] |
| | 229 | 29.3% | 60.1% | 8.9% | Pena et al., 2011[149] |
| **Center-West** | 1,053 | 24.2% | 62.7% | 13.1% | de Souza et al., 2019[50] |
| | 126 | 26% | 58% | 16% | Saloum et al., 2013[148] |
| **Southeast** | 1,442 | 78.5% | 14.7% | 6.7% | Kehdy et al., 2015[150] |
| | 8,791 | 19.2% | 72.3% | 7.6% | de Souza et al., 2019[50] |
| | 509 | 27% | 61% | 12% | Saloum et al., 2013[148] |
| | 264 | 17.3% | 74.2% | 7.3% | Pena et al., 2011[149] |
| **South** | 3,736 | 15.9% | 76.1% | 8% | Kehdy et al., 2015[150] |
| | 11,078 | 8.4% | 81.8% | 8.6% | de Souza et al., 2019[50] |
| | 64 | 15% | 74% | 11% | Saloum et al., 2013[148] |
| | 238 | 10.3% | 79.5% | 9.4% | Pena et al., 2011[149] |
| **Brazil** | 31,851 | 19.6% | 68.1% | 11.6% | de Souza et al., 2019[50] |
| | 1,304 | 23% | 60% | 17% | Gontijo et al., 2018[151] |
| | 934 | 16.8% | 70.6% | 11.1% | Pena et al., 2011[149] |

**Table S3. Ancestry proportions estimated among quilombo remnants populations.** Ancestry estimates for quilombo remnant populations, identified by Brazilian state (SE - Sergipe; RS - Rio Grande do Sul; PA - Pará; GO - Goiás; BA - Bahia; RO - Rondônia; AM - Amazonas; SP - São Paulo), along with sample size, ancestry proportions, and the study source. "Amazônia / AM" and "Vale do Ribeira / SP" represent multiple communities.

| Population/State | Sample size (N) | Ancestry Proportions | | | Study Source |
|---|---|---|---|---|---|
| | | African | European | Native American | |
| **Mocambo/SE** | 81 | 42.6% | 44.9% | 12.2% | Gontijo et al., 2018[151] |
| **Paredão/RS** | 32 | 80.2% | 19.8% | 0.0% | Bortolini et al., 1995[152] |
| **Cametá/PA** | 94 | 48.0% | 17.9% | 34.1% | Bortolini et al., 1995[152] |
| **Trombetas/PA** | 191 | 56.4% | 23.8% | 19.8% | Bortolini et al., 1995[152] |
| **Sacutiaba/BA** | 30 | 43.0% | 46.8% | 7.40% | Gontijo et al., 2018[151] |
| **Kalunga/GO** | 72 | 67.3% | 24.9% | 7.20% | Gontijo et al., 2018[151] |
| **Santiago do Iguape/BA** | 37 | 56.8% | 2.5% | 40.7% | Gontijo et al., 2014[153] |
| **Santo Antônio do Guaporé/RO** | 31 | 37.6% | 20.4% | 42.0% | Gontijo et al., 2014[153] |
| **Amazônia/AM (7 comm.)** | 294 | 48.5% | 28.9% | 22.6% | Maciel et al., 2011[154] |
| **Vale do Ribeira/SP (10 comm.)** | 307 | 39.7% | 39.0% | 21.3% | Kimura et al., 2013[59] |
| **Vale do Ribeira/SP (8 comm.)** | 431 | 47.4% | 36.3% | 16.1% | Present Study |

**Table S4. Summary of Regions of Interest (ROIs).** The table lists all the ROIs identified during the pedigree analysis, including those that were ultimately discarded (due to absence of corroboration between the subpanels). Columns include Pedigree, Peak LOD Score, rsID interval, Genetic Location (cM), and Physical Positions in both hg19 and hg38 builds.

| Final ROIs | Pedigree | Peak LOD Score | rsID interval | Genetic Location (cM) | Phys. Position hg19 (BP) | Phys. Position hg38 (BP) |
|---|---|---|---|---|---|---|
| **1** | TU | 1.503 | rs454510-rs7536191 | 149.22-154.58 | 1:120195042-152783560 | 1:119652419-152811084 |
| **2** | ABDR | 1.824 | rs1009591-rs804134 | 25.89-33.38 | 1:11928830-15150914 | 1:11868773-14824418 |
| **3** | ABDR | 2.161 | rs61834059-rs6695129 | 252.09-256.37 | 1:236655728-237993416 | 1:236492428-237830116 |
| **4** | PC | 1.9063 | rs6546079-rs905489 | 85.11-89.42 | 2:64725466-68111009 | 2:64498332-67883877 |
| **5** | ANNH | 3.0363 | rs6798365-rs9863587 | 196.56-223.78 | 3:186177610-197581147 | 3:186459821-197854276 |
| **6** | IV | 1.9381 | rs11133180-rs12507909 | 50.97-75.08 | 4:32130034-58698209 | 4:32128412-57832043 |
| **7** | IV | 2.3224 | rs7658883-rs28611030 | 169.14-206.59 | 4:166837654-187448441 | 4:165916502-186527287 |
| **8** | TU | 2.1174 | rs10078603-rs6867772 | 73.13-81.13 | 5:56865792 - 67963112 | 5:57569965-68667285 |
| **9** | TU | 2.6129 | rs17697699-rs558825 | 29.91-42.98 | 6:12027402-20046786 | 6:12027169-20046555 |
| **10** | ABDR | 2.65 | rs9969455-rs34268501 | 24.86-42.25 | 8:12617155-22460204 | 8:12759646-22602691 |
| **11** | IV | 1.4158 | rs7043980-rs7044521 | 158.86-162.55 | 9:137876807-138699102 | 9:134984961-135807256 |
| **12** | ANNH | 2.7959 | rs1774718-rs10884028 | 112.51-122.38 | 10:95166828-106396917 | 10:93407071-104637159 |
| **13** | GASP | 2.414 | rs894108-rs651861 | 81.83-88.91 | 11:71082971-78681489 | 11:71371925-78970444 |
| **14** | IV | 2.662 | rs1981751-rs306656 | 37.36-52.47 | 12:16538591-27693531 | 12:16385657-27540598 |
| **15** | IV | 2.087 | rs10431335-rs10870508 | 158.98-175.66 | 12:128679068-133341344 | 12:128194523-132764758 |
| **16** | GASP | 2.5548 | rs2146878-rs9318029 | 49.54-64.81 | 13:46724113-72365249 | 13:46149978-71791117 |
| **17** | TU | 2.1751 | rs12446759-rs7191751 | 109.42-126.19 | 16:81773003-86304261 | 16:81739398-86270655 |
| **18** | IV | 1.771 | rs11651877-rs4796444 | 7.00-17.37 | 17:1852831-6135803 | 17:1949537-6232483 |
| **19** | IV | 2.1387 | rs7208501-rs1696757 | 90.46-129.42 | 17:58912708-77800743 | 17:60835347-79826944 |
| **20** | TU | 1.6084 | rs56757480-rs9967057 | 83.85-88.41 | 18:56412371-58117122 | 18:58745139-60449889 |
| **21** | ANNH | 2.503 | rs4805131-rs7252448 | 61.27-72.29 | 19:36097297-46142053 | 19:35606395-45638795 |
| **22** | ANNH | 1.86 | rs273646-rs55747023 | 86.95-100.08 | 19:51752059-54875458 | 19:51248804-54363851 |
| **discarded** | ANNH | 1.3101 | rs6532990-rs2672477 | 112.40-116.30 | 4:103013903-108498781 | 4:102092746-107577624 |
| **discarded** | ANNH | 1.4651 | rs2575632-rs7440784 | 117.43-124.55 | 4:109586178-116434752 | 4:108665022-115513596 |
| **discarded** | ANNH | 1.3455 | rs34124147-rs4404544 | 120.64-122.27 | 4:112231048-113858241 | 4:111309892-112937085 |
| **discarded** | TU | 1.5337 | rs36088-rs28575551 | 75.95-83.44 | 5:60930305-71799312 | 5:61634478-72503485 |
| **discarded** | PC | 1.7203 | rs62521114-rs7840329 | 78.68-94.25 | 8:65637942-81802855 | 8:64725385-80890620 |
| **discarded** | ANNH | 1.7482 | rs861136-rs1049632 | 31.91-40.16 | 10:13318060-16796919 | 10:13276060-16754920 |
| **discarded** | PC | 1.4896 | rs1709763-rs929693 | 57.05- 59.79 | 16:27897258-47986157 | 16:27885937-47952246 |
| **discarded** | ANNH | 2.368 | rs7251550-rs6510896 | 3.46-18.93 | 19:1998286-6402648 | 19:1998287-6402637 |

**Table S6. List of 117 suggestive and/or significantly associated variants.** Where "pop." is population-based and "ped" is pedigree-based imputation strategy. For each variant, the identifier (rsID), variant physical position, gene name and variant consequence are described (where "WE" is without effect, "N.A." is not available, and "CTCF" is CTCF binding site). Variants are listed based on ROIs order. The red asterisk (*) indicates variants found in genes in common with EH-genes investigation.

| ROIs | Pop./Ped. Based | SNP (rsID) | Location (GRCh37) | Gene | Consequence |
|---|---|---|---|---|---|
| **1** | Pop. | rs10888437 | 1:151,809,238 | intergenic | Regulatory CTCF/ promoter |
| **1** | | rs539304* | 1:120,275,602 | *PHGDH* | Intron/WE |
| **1** | | rs539426* | 1:120,275,647 | *PHGDH* | Intron/WE |
| **1** | Ped. | rs59921678 | 1:147,387,781 | intergenic | N/A |
| **1** | | rs10494229* | 1:120,229,117 | *PHGDH* | Intron/WE |
| **1** | | rs11577560* | 1:120,229,865 | *PHGDH* | Intron/WE |
| **1** | | rs78976434* | 1:151,965,380 | *S100A10* | Intron/WE |
| **1** | | rs80184297 | 1:151,970,608 | *NBPF18P* | Intron/WE |
| **1** | | rs187215567 | 1:151,971,216 | *NXPE2/NXPE4* | Intron/WE |
| **1** | | rs4845725 | 1:151,973,800 | *NBPF18P* | Intron/WE |
| **2** | Pop. | rs4845892* | 1:12,073,785 | *MFN2* | Regulatory/promoter |
| **2** | Ped. | rs72875205 | 1:14,338,444 | *KAZN* | Intron/WE |
| **2** | | rs3010942 | 1:12,740,964 | intergenic | N/A |
| **3** | Pop. | rs6678561* | 1:237,982,847 | *RYR2* | Intron/WE |
| **3** | | rs4659809* | 1:237,983,213 | *RYR2* | Intron/WE |
| **3** | | rs4659808* | 1:237,983,074 | *RYR2* | Intron/WE |
| **3** | | rs10925518* | 1:237,979,815 | *RYR2* | Intron/WE |
| **3** | | rs10925517* | 1:237,979,806 | *RYR2* | Intron/WE |
| **3** | | rs1558491940* | 1:237,987,843 | *RYR2* | Intron/WE |
| **3** | | rs10802631* | 1:237,860,401 | *RYR2* | Intron/WE |
| **3** | | rs12022241* | 1:237,656,124 | *RYR2* | Intron/WE |
| **3** | | rs575448073* | 1:237,656,181 | *RYR2* | Intron/WE |
| **3** | Ped. | rs2564723 | 1:236,791,038 | intergenic | N/A |
| **3** | | rs646040* | 1:236,606,292 | *EDARADD* | Intron/WE |
| **3** | | rs12117452* | 1:237,647,282 | *RYR2* | Intron/WE |
| **3** | | rs10925261* | 1:237,053,186 | *MTR* | Intron/WE |
| **3** | | rs41413447 | 1:236,659,905 | intergenic | N/A |
| **3** | | rs6671704 | 1:236,797,237 | intergenic | N/A |
| **3** | | rs56042629 | 1:236,797,854 | intergenic | N/A |
| **3** | | rs1805087* | 1:237,048,500 | *MTR* | Missense/LB |
| **4** | Ped. | rs2160390 | 2:65,855,537 | *LINC02934* | Intron/WE |
| **4** | | rs2422278* | 2:64,879,668 | *SERTAD2* | Intron/Regulatory Promoter |
| **4** | | rs4308115 | 2:68,028,751 | *LINC01812* | Intron/WE |
| **4** | | rs17032545 | 2:67,017,450 | intergenic | N.A. |
| **4** | | rs7563552 | 2:68,029,651 | *LINC01812* | Intron/WE |
| **4** | | rs7602687 | 2:68,029,703 | *LINC01812* | Intron/WE |
| **5** | Pop. | rs76929915 | 3:195,849,315 | intergenic | N.A. |
| **5** | | rs73055915 | 3:190,144,799 | *TMEM207* | 3'UTR/WE |
| **5** | | rs11917973 | 3:195,838,613 | intergenic | N.A. |
| **5** | | rs73055924 | 3:190,148,009 | *TMEM207* | Intron/WE |
| **5** | | rs58109815 | 3:190,125,215 | *CLDN16* | Intron/WE |
| **5** | | rs200021867 | 3:190,640,882 | intergenic | N.A. |
| **5** | | rs190184306 | 3:190,640,875 | intergenic | N.A. |
| **5** | | rs201379805 | 3:190,640,881 | intergenic | N.A. |

| | | | | | |
|---|---|---|---|---|---|
| **5** | | rs978941* | 3:188,271,942 | *LPP* | Intron/Regulatory Promoter |
| **5** | Ped. | rs9990333 | 3:195,827,205 | intergenic | N.A. |
| **5** | | rs73182261 | 3:186,134,843 | intergenic | N.A. |
| **5** | | rs555412 | 3:195,804,918 | *TFRC* | Intron/WE |
| **5** | | rs6794037 | 3:194,789,163 | *XXYLT1* | 3'UTR/WE |
| **5** | | rs6797948 | 3:194,784,705 | *XXYLT1* | 3'UTR/Regulatory CTCF/Promoter |
| **7** | Pop. | rs58367421 | 4:167,016,808 | *TLL1* | Intron/WE |
| **7** | | rs76416003 | 4:181,942,978 | intergenic | N.A. |
| **7** | | rs17047248 | 4:167,011,894 | *TLL1* | Intron/WE |
| **7** | | rs77213883 | 4:167,012,672 | *TLL1* | Intron/Regulatory CTCF |
| **7** | | rs4861699 | 4:186,984,740 | intergenic | Intron/WE |
| **7** | Ped. | rs6552571 | 4:169,175,897 | DDX60 | Intron/WE |
| **7** | | rs10009131 | 4:183,391,291 | *TENM3* | Intron/WE |
| **8** | Ped. | rs73769903 | 5:67,650,229 | intergenic | Regulatory CTCF/promoter |
| **8** | | rs10471750 | 5:67,737,628 | intergenic | Regulatory/promoter |
| **9** | Pop. | rs114362954 | 6:14,913,620 | intergenic | N.A. |
| **9** | | rs11962715 | 6:16,809,219 | intergenic | N.A. |
| **9** | | rs147280918 | 6:19,982,505 | intergenic | N.A. |
| **9** | | rs112980988 | 6:14,958,938 | intergenic | Regulatory/promoter |
| **9** | Ped. | rs980959 | 6:18,733,216 | intergenic | Regulatory/open cromatin |
| **9** | | rs12216318 | 6:12,409,416 | intergenic | Regulatory/promoter |
| **10** | Pop. | rs7836144 | 8:19,625,912 | intergenic | N.A. |
| **10** | | rs7837390 | 8:19,626,120 | intergenic | N.A. |
| **10** | | rs4922096 | 8:19,621,954 | intergenic | N.A. |
| **10** | | rs112999507 | 8:19,621,501 | intergenic | N.A. |
| **10** | | rs6651482 | 8:19,622,598 | intergenic | N.A. |
| **10** | | rs10088110 | 8:19,622,753 | intergenic | Regulatory/Open cromatin |
| **10** | | rs10088203 | 8:19,622,809 | intergenic | Regulatory/Open cromatin |
| **10** | | rs11782067 | 8:19,623,129 | intergenic | Regulatory/Open cromatin |
| **10** | | rs10087365 | 8:19,622,859 | intergenic | Regulatory/Open cromatin |
| **10** | | rs6651481 | 8:19,622,349 | intergenic | N.A. |
| **10** | | rs35045637 | 8:18,675,486 | *PSD3* | Intron/WE |
| **10** | | rs1585175565 | 8:20,472,294 | *SNORD3F* | 3'UTR/WE |
| **10** | Ped. | rs6651387 | 8:15,119,655 | intergenic | N.A. |
| **11** | Ped. | rs2031825 | 9:138,026,999 | intergenic | N.A. |
| **11** | | rs7037014* | 9:138,643,920 | *KCNT1* | Intron/WE |
| **11** | | rs1165017* | 9:138,602,228 | *KCNT1* | Intron/WE |
| **12** | Pop. | rs2038563 | 10:101,413,863 | *SLC25A28* | Intron/WE |
| **13** | Ped. | rs3819242 | 11:74,407,676 | *CHRDL2* | Intron/Regulatory Promoter |
| **13** | | rs1435468* | 11:78,744,472 | *TENM4* | Intron/WE |
| **13** | | rs10793116 | 11:74,920,005 | *TPBGL-AS1* | Intron/WE |
| **14** | Ped. | rs59714345 | 12:26,135,679 | *RASSF8* | Intron/Regulatory Enhancer |
| **15** | Ped. | rs61934871 | 12:132,102,428 | intergenic | N.A. |

| | | | | | |
|---|---|---|---|---|---|
| **18** | Pop. | rs9899953* | 17:3,809,282 | *P2RX1* | Intron/Regulatory Promoter |
| **18** | | rs2585266 | 17:5,213,391 | *RABEP1* | Intron/WE |
| **18** | | rs28484653 | 17:5,976,351 | *WSCD1* | Intron/WE |
| **18** | | rs4796306 | 17:5,978,414 | *WSCD1* | Intron/WE |
| **18** | | rs8065049 | 17:5,967,113 | *WSCD1* | Intron/WE |
| **18** | | rs542170600* | 17:3,993,829 | *ZZEF1* | Intron/WE |
| **18** | Ped. | rs9909751 | 17:4,299,256 | intergenic | Regulatory/promoter |
| **18** | | rs7220105 | 17:3,878,021 | intergenic | N.A. |
| **18** | | rs9082* | 17:1,801,263 | *RPA1* | 3' UTR/WE |
| **19** | Pop. | rs12232542 | 17:73,056,757 | *KCTD2* | Intron/Regulatory Promoter |
| **19** | | rs72631377 | 17:73,053,608 | *KCTD2* | Intron/WE |
| **19** | | rs112276578 | 17:73,052,008 | *KCTD2* | Intron/WE |
| **19** | | rs1000353180 | 17:73,058,659 | *KCTD2* | Intron/WE |
| **19** | | rs12602540 | 17:73,060,111 | *KCTD2* | 3'UTR/WE |
| **19** | | rs56284738 | 17:73,062,836 | *KCTD2* | 3'UTR/Regulatory Promoter |
| **19** | | rs201766965 | 17:73,054,433 | *KCTD2* | Intron/WE |
| **19** | | rs12943148 | 17:73,051,732 | *KCTD2* | Intron/WE |
| **19** | | rs12936866 | 17:73,052,050 | *KCTD2* | Intron/WE |
| **19** | | rs57813751 | 17:73,053,512 | *KCTD2* | Intron/WE |
| **19** | | rs56267297 | 17:73,046,996 | *KCTD2* | Intron/WE |
| **19** | | rs113775187 | 17:73,047,290 | *KCTD2* | Intron/WE |
| **19** | | rs11871228 | 17:73,047,642 | *KCTD2* | Intron/WE |
| **19** | Ped. | rs16974626 | 17:67,925,806 | intergenic | N.A. |
| **19** | | rs9909931 | 17:74,368,088 | *PRPSAP1* | Intron/Regulatory Promoter |
| **20** | Ped. | rs11665647* | 18:56,247,976 | *ALPK2* | Intron/Regulatory CTCF |
| **20** | | rs12457935* | 18:56,248,929 | *ALPK2* | Intron/WE |
| **21** | Pop. | rs57339845 | 19:44,279,789 | *KCNN4* | Intron/WE |
| **21** | Ped. | rs80232770 | 19:44,383,569 | *ZNF404* | Intron/WE |
| **21** | | rs16976860 | 19:44,358,761 | intergenic | N/A |
| **22** | Ped. | rs1050527 | 19:54,677,167 | *MBOAT7* | 3'UTR/Regulatory Promoter |

**Table S7. Linkage disequilibrium (LD) results for missense variants.** Variant 1 represents variants suggestive/significantly associated with the trait, while Variant 2 indicates variants in linkage disequilibrium with Variant 1. The table includes variant locations, LD metrics ($r^2$ and D'), and populations from the 1000 Genomes Project (1000G. POP.), showing strong LD ($r^2$ and D' close to or equal to 1) across different populations, including CEU (Utah residents with Northern and Western European ancestry), YRI (Yoruba in Ibadan, Nigeria), CLM (Colombians in Medellín), MXL (Mexican ancestry in Los Angeles), and PEL (Peruvians from Lima).

| Variant 1 | Variant 1 location | Variant 2 | Variant 2 location | $r^2$ | D' | 1000G. POP. |
|---|---|---|---|---|---|---|
| **rs59921678** | 1:147387781 | rs142935665 | 1:147415585 | 1 | 1 | CEU |
| **rs10925261** | 1:237053186 | rs1805087 | 1:237048500 | 1 | 1 | CEU |
| **rs1050527** | 19:54677167 | rs77215230 | 19:54675643 | 0.951 | 1 | CEU |
| **rs555412** | 3:195804918 | rs3817672 | 3:195800811 | 0.839 | 0.999 | YRI |
| **rs80232770** | 19:44383569 | rs77337446 | 19:44378224 | 1 | 1 | YRI |
| **rs58109815** | 3:190125215 | rs74418158 | 3:190374230 | 1 | 1 | CLM |
| **rs80232770** | 19:44383569 | rs77779894 | 19:44378178 | 1 | 1 | CLM |
| **rs80232770** | 19:44383569 | rs76550420 | 19:44377048 | 1 | 1 | CLM |
| **rs6671704** | 1:236797237 | rs2243525 | 1:236706862 | 0.723 | 0.906 | MXL |
| **rs6671704** | 1:236797237 | rs2275689 | 1:236721660 | 0.723 | 0.906 | MXL |
| **rs11665647** | 18:56247976 | rs9944810 | 18:56247600 | 0.778 | 0.999 | MXL |
| **rs10793116** | 11:74920005 | rs140226575 | 11:74978732 | 0.701 | 0.999 | PEL |
| **rs80232770** | 19:44383569 | rs59926292 | 19:44536071 | 1 | 1 | PEL |
| **rs80232770** | 19:44383569 | rs138492321 | 19:44153248 | 1 | 1 | PEL |
| **rs80232770** | 19:44383569 | rs11880330 | 19:44535975 | 1 | 1 | PEL |
| **rs59921678** | 1:147387781 | rs142935665 | 1:147415585 | 1 | 1 | CEU |
| **rs10925261** | 1:237053186 | rs1805087 | 1:237048500 | 1 | 1 | CEU |
| **rs1050527** | 19:54677167 | rs77215230 | 19:54675643 | 0.951 | 1 | CEU |

**Table S8. Weighted scores for ROIs ranking and prioritization.** A) EH genes refer to the occurrence, or not, of genes related to EH in the literature; B) Linkage analysis (LA) scored by peak LOD score and consensus between subpanels; C) Association studies (AS) scored based on qualitatively interpreted p-value results in relation to the threshold; D) Relevance of the outcome based the extent to which each gene has the potential to explain the phenotype.

| EH Genes* | Linkage Analysis | | Association Studies Sugg./ Signif. | Gene Relevance** |
|---|---|---|---|---|
| | Peak LOD Score | Subpanels consensus | | |
| Yes: 1 point | 1.40-1.69: 1 point | 1 subpanel: 1 point | Below sugg.: 0 point | Very low: 1 point |
| No: 0 point | 1.70-1.99: 2 points | 2 subpanels: 2 points | Slight above sugg.: 1 point | Low: 2 points |
| | 2.00-2.29: 3 points | 3 subpanels: 3 points | Above sugg.: 2 points | Medium: 3 points |
| | 2.30-2.59: 4 points | | Above signif.: 3 points | Above-medium: 4 points |
| | 2.60-2.89: 5 points | | | High: 5 points |
| | 2.90-3.19: 6 points | | | |

*Genes already referenced in the literature as EH-related.
**Arbitrary score based on the gene potential to explain the phenotype (as biological function, corroborating studies, presence of SNPs associated with EH).
